# Policy approaches to decarbonising the transport sector in Aotearoa/NZ: Equity, health and health system impacts

**DOI:** 10.1101/2024.01.29.24301894

**Authors:** Caroline Shaw, Anja Mizdrak, Ryan Gage, Melissa McLeod, Rhys Jones, Alistair Woodward, Linda Cobiac

## Abstract

**Background:** Health co-benefits are a key potential advantage of transport decarbonisation policy. However, health impacts will occur in the context of existing transport-health inequities and decarbonisation policies will themselves impact inequities.

**Methods:** We modelled the health, health system and environmental impacts of the ‘Behaviour’ and ‘Technology’ pathways developed by the New Zealand Climate Change Commission. Household transport related health impacts were modelled through the pathways of physical activity, air pollution (PM_2·5_ and NO_2_) and injury for the New Zealand population from 2018 to 2050. We modelled impacts for Māori (the Indigenous Peoples of Aotearoa) and non-Māori.

**Findings:** Both pathways show improvements in population health, reductions in health system costs and reduced lifecycle greenhouse gas emissions compared to baseline, although health gains were substantially larger in the Behaviour pathway. Health gains were 20-30% larger for Māori than non-Māori in both pathways, although more healthy life years were gained by Māori in the Behaviour pathway. For those aged 0-4 in 2018, healthy life expectancy differences between Māori and non-Māori reduced by 0·5% in the Behaviour pathway. Healthy life years gained by Māori and non-Māori altered substantially depending on assumptions about the equity of the implemented pathway.

**Interpretation:** Decarbonising transport may reduce health and healthy life expectancy inequities between Māori and non-Māori if policies supporting decarbonisation are implemented equitably. Pathways that increase physical activity will have a much larger impact on population health than those which rely on low emission vehicles. **Funding:** Health Research Council of New Zealand (20/151) and University of Otago.

## Introduction

Health co-benefits, particularly in the near term, are one of the key advantages of transport decarbonisation policy.^1^ Current transport systems are a significant source of death and disability through multiple pathways.^2,3^ Modelling studies mostly confirm that scenarios to decarbonise transport, or specific policies that might be part of decarbonisation scenarios, would have health benefits.^4-7^ However, the magnitude of the modelled health benefits depend on range of factors including the scenario or policy mix, local context and model related factors.

Current transport systems create and perpetuate substantial health (and other) inequities.^2,8^ Broadly, less socially advantaged groups are more likely to be excluded from the benefits of the current system and disproportionately experience the health harms from noise exposure, air pollution, physical inactivity, injury and climate change.^2,9^ Conversely, more advantaged groups systematically drive and fly more, thus causing more environmental and health damage.^10^ Transport decarbonisation policy will be overlaid on these pre-existing inequities. While decarbonisation has been identified as an opportunity to reduce inequities,^11^ relatively little research has focused on how proposed decarbonisation scenarios will impact on transport-health inequities.^12^

In Aotearoa/New Zealand (hereafter Aotearoa) the current, car-dominated transport system has been estimated to cause as much ill-health each year as tobacco or obesity through the pathways of air pollution, physical inactivity and injury.^13^ This is likely to be a significant underestimate, as important pathways between transport and health were not included in this estimate (e.g. NO_2_). In addition, the health inequities from transport contribute at least 2-3% of the 7-8 year difference in healthy life expectancy between Māori, the Indigenous people of Aotearoa, and non-Māori.^13^

Transport decarbonisation policy has developed rapidly in Aotearoa in recent years. The 2019 Climate Change Response (Zero Carbon) Amendment Act resulted in the creation of an independent organisation, He Pou a Rangi/Climate Change Commission (CCC). In 2021 the CCC established carbon budgets and outlined a range of possible decarbonisation pathways for all emitting sectors, including transport.^14^ While health co-benefits were discussed by the CCC, no attempt was made to quantify them.

Given the importance of transport to health and health inequity and the opportunity afforded by decarbonisation policy, this paper examines the impact of transport decarbonisation pathways proposed by the CCC on population health, health inequity between Māori and non-Māori, and the health system in Aotearoa.

## Methods

### Decarbonisation pathways

The CCC developed seven national level pathways to net zero emissions by 2050 for the transport sector.^14^ In this paper we simulate health outcomes for the two pathways that were most distinct. Compared to current patterns, the CCC named “Further behaviour change” pathway emphasises increased levels of cycling, public transport use, and vehicle kilometres travelled (VKT) reduction alongside light fleet electrification. The CCC’s “Further technology change” pathway focuses on high uptake of electric vehicle technology, with more modest increases in public transport use and reductions in VKT. We compared these decarbonisation pathways (henceforth referred to as ‘Behaviour’ and ‘Technology’ for brevity) with a counterfactual business-as-usual pathway (BAU), where we assume a continuation of current (2018) household travel patterns and light fleet composition. Figure 1 illustrates key aspects of the pathways. Baseline mode-specific VKT was based on the NZ Household Travel Survey 2015-2018, which illustrates the high levels of car dependency in Aotearoa. The annual percentage changes in VKT for each CCC pathway were based on the projected increases estimated by the CCC. Further details on the process of aligning the CCC pathways with this model are in the supplementary information (SI).

**Figure 1.**
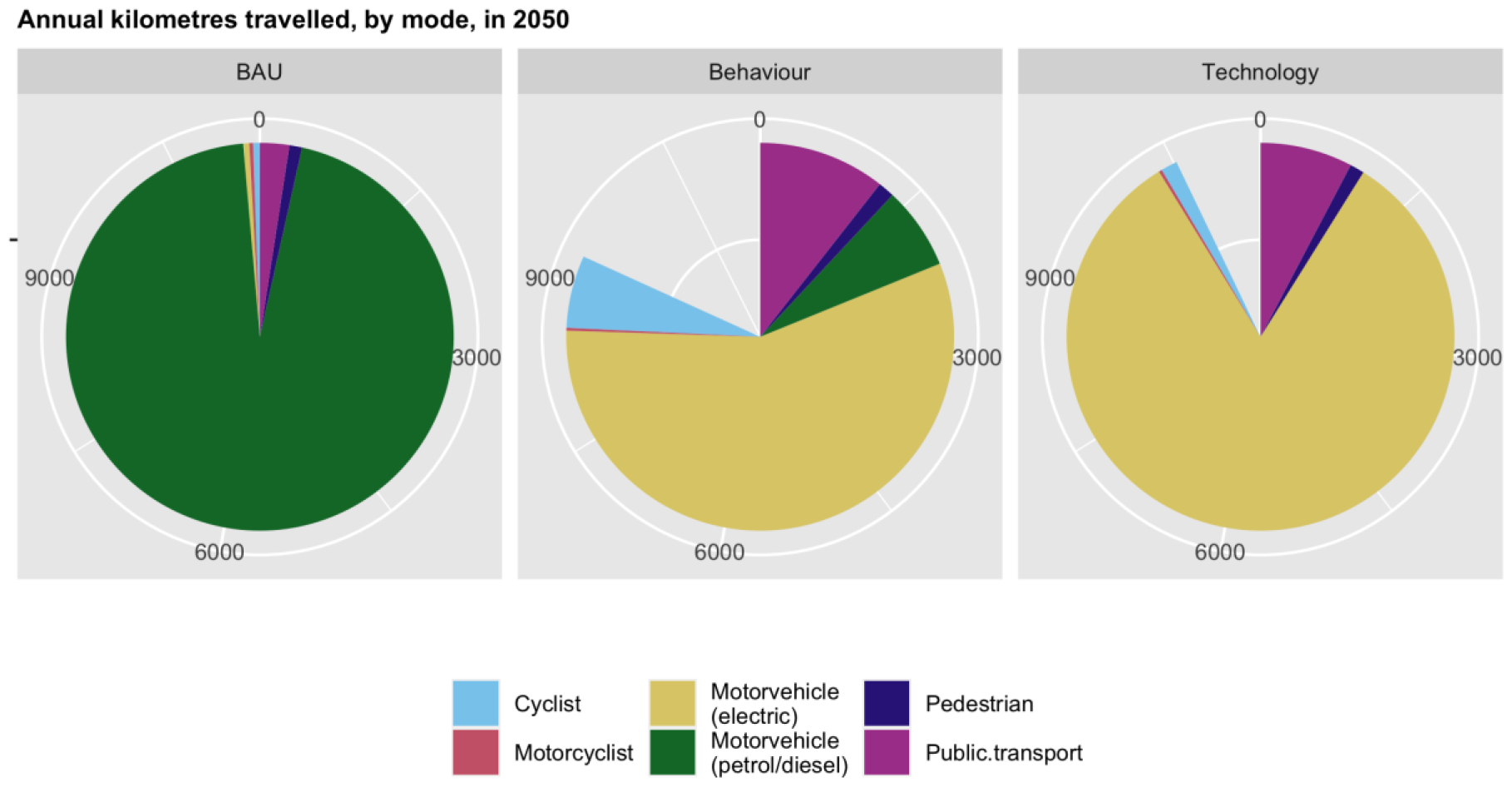
Annual km travelled by mode in 2050 for business-as-usual (BAU), Further Behaviour Change (‘Behaviour’) and Further Technology Change (‘Technology’) pathways Note: greyed out areas of the pie chart represent reductions in VKT compared to BAU.

### Model

We simulated the impact of the CCC’s two decarbonisation pathways for household land transport (i.e. the model excludes commercial and air transport) on population health (via physical activity, air pollution and injuries), greenhouse gas emissions (GHGe), health inequities and health system costs in Aotearoa using a model that we developed in R (Version 4·2·2).^15^ A diagrammatic overview of the model and further information on methods are available in the SI.

We started the model in 2018 with the Aotearoa population, defined by 5-year age group, gender (male/female) and ethnicity (Māori/non-Māori). Each year, we calculated changes in transport patterns and health outcomes. A new cohort of 0-4 years olds entered the model every five years of the simulation, based on projected population growth. We ran the model until 2118 (100 years) with cohorts reducing in size per year in accordance with their background mortality rate or being censored at age 100 years. We assumed that transport patterns for the scenarios would stabilise beyond 2050.

The CCC makes no predictions about how the changes in transport patterns are distributed across the population by age, gender or ethnicity. In our simulations we assumed that the current age patterns of transport, which are highly related to life-stage (e.g., school, work, retirement), would remain proportionally the same in the future. However, we considered different equity scenarios around, firstly, the distribution of predicted travel changes by gender and ethnicity and, secondly, equity in background health (see Table 1).

**Table 1.** Modelled equity scenarios for each CCC decarbonisation pathway.

| Equity scenario | Description |
| --- | --- |
| **Equity in implementation** |  |
| Base case | Changes in travel distributed equally across males and females, Māori and non-Māori (i.e., everyone makes the same absolute VKT per capita changes in travel, regardless of gender or ethnicity). |
| Proportional impact | Changes in travel modelled as a percentage change to current participation by mode, i.e., the same percentage change will produce different absolute change by age, ethnicity and gender due to differing underlying rates |
| Equitable effect | Changes in travel distributed to achieve equal participation within each travel mode, by gender and ethnicity, after 10 years, i.e. eradicating current differences in transport by gender and ethnicity in all modes.<sup>16</sup> |
| Cycling equity | Changes in travel distributed to achieve equal cycling participation for all groups, at a threshold cycling mode share of 7%.<sup>17</sup> |
| **Equity in background health** |  |
| Equal mortality | Māori background rates of mortality are substituted with non-Māori background mortality rates. |
| Equal morbidity | Māori background rates of morbidity are substituted with non-Māori background morbidity rates. |
| Equal mortality and morbidity | Māori background rates of mortality and morbidity are substituted with non-Māori background mortality and morbidity rates. |

#### Health pathways

Air pollution: We estimated changes in PM_2·5_ and NO_2_ from projected changes in distance travelled by mode and the proportion of fleet electrification. We based tailpipe and other (e.g. road wear) emissions per kilometre travelled by car, motorcycle and bus, for each fuel source (petrol, diesel, electric, hybrid, or plug-in hybrid) on New Zealand Transport Agency data.^18,19^ We modelled the impact of transport-related changes in air pollution on ischaemic heart disease (PM_2·5_), stroke (PM_2·5_) and asthma (NO_2_) using relative risks from a recent NZ population-wide cohort study.^20^

Physical activity: We simulated the impact of physical activity changes onto the incidence of cardiovascular diseases, diabetes, various cancers and depression using relative risks from systematic reviews of prospective cohort studies.^21,22^ Baseline non-occupational physical activity levels were ascertained from population surveys,^23,24^ and changes in physical activity were determined from the projected changes in distance travelled by walking and cycling modes. We assumed METS (metabolic equivalents) of the following active modes: walking 2·0 METS, conventional cycling 5·8METS, ebiking 4·97METS, escootering 1·5 METS (all adjusted for baseline metabolic rate).

BMI: We additionally calculated the impact of changes in physical activity on BMI, although we did not quantify flow-on effects on obesity-related diseases to avoid double-counting of health impacts from physical activity. To estimate changes in BMI, we first estimated changes in energy expenditure from physical activity associated with active transport, accounting for variations in resting metabolic rate by age, gender and ethnicity,^23^ and assuming a food compensation factor of 0·57 (95% uncertainty interval: 0·19, 0·96).^25^ We derived body mass index (BMI) effects from the changes in energy expenditure using a time-lagged dose response of 94 (95% CI: 88·2, 99·8) kJ/kg/day.^26^ Changes in BMI were presented as prevalence of overweight (BMI between 25 and 30) and obesity (BMI > 30).

Transport injuries: We simulated the impact of changing travel patterns on risk of cyclist, pedestrian, motor vehicle and motor cyclist road injuries, using hospitalisation data to separate mode-specific rates of fatal and non-fatal injuries by victim and striker (if any). We applied ‘safety-in-numbers’ coefficients^27^ to reflect the non-linear relationship between mode volume and injury risk.

## Outputs

Population health: We simulated changing incidence of disease and injury over time using a proportional multi-state lifetable approach·4^5^ The health gain associated with each decarbonisation pathway was calculated in health-adjusted life years (HALYs) from the difference in years of life lived by the population, adjusted for time spent in ill-health using morbidity rates derived from New Zealand and Global Burden of Disease study disability weights.^28,29^ Health-adjusted life expectancy (HALE) was calculated from the difference in health-adjusted years of life lived by each age group in compared to the BAU population.

Health equity: We estimated impact on health equity by age-standardising (using the 2001 Census total Māori population^30^) the lifetime HALY rate for the baseline population and calculating the Māori/non-Māori rate ratio. Since higher background rates of mortality and morbidity may (unfairly) limit the magnitude of modelled health gains for Māori, we ran three additional equity scenarios (see Table 1) where we substituted Māori with non-Māori rates for: background mortality, background morbidity, and both mortality and morbidity.^31^

Health system costs: We estimated the impact of the decarbonisation pathways on costs to the Aotearoa/New Zealand health system from: (a) the modelled difference in number of cases of each transport-related cause and the unit costs of treatment; and (b) the modelled difference in number of people alive and the average cost of health care for all other causes. Unit costs were derived using published methods from models fitted to nationally linked health data for New Zealand adults for the financial years 2015/16 to 2017/2018.^32^

Greenhouse gas emissions: We simulated changes in lifecycle CO_2_ equivalents for each pathway as a result of changes in light fleet composition (primarily electrification) and changes in VKT and mode distribution. We included tailpipe emissions, other vehicle emissions (related to manufacturing and cycling, operational and infrastructure) and food related emissions due to extra energy required for walking and cycling. Most emissions factors were sourced from local data,^18^ supplemented with literature, bespoke calculations or assumptions as required.

Uncertainty analysis: We evaluated means and 95% uncertainty intervals for all modelled outputs using Monte Carlo analysis (2000 iterations). All values are rounded to two significant figures.

Sensitivity analyses: We tested the pathways using different assumptions around key parameters, including lower electric car uptake (75% of light fleet by 2050^33^), lower cycling injury rates (96% reduction in incidence and 93% reduction in mortality-more akin to the Netherlands^34^) and a less GHGe intensive diet (30% reduction^35^).

Funding for this project was received from the Health Research Council of New Zealand (grant number 20/151) and University of Otago. The funders had no role in the project. Ethics approval was granted by the University of Otago Ethics Committee (HD20/081).

## Results

### Overall impact

Both the Behaviour and Technology modelled pathways will improve population health and avert health system costs when compared to business as usual. Māori males and females gain more HALYs per 1000 than non-Māori in both decarbonisation pathways under the base case assumption that everyone can make the same changes to travel, regardless of gender or ethnicity (Table 3).

**Table 3.**
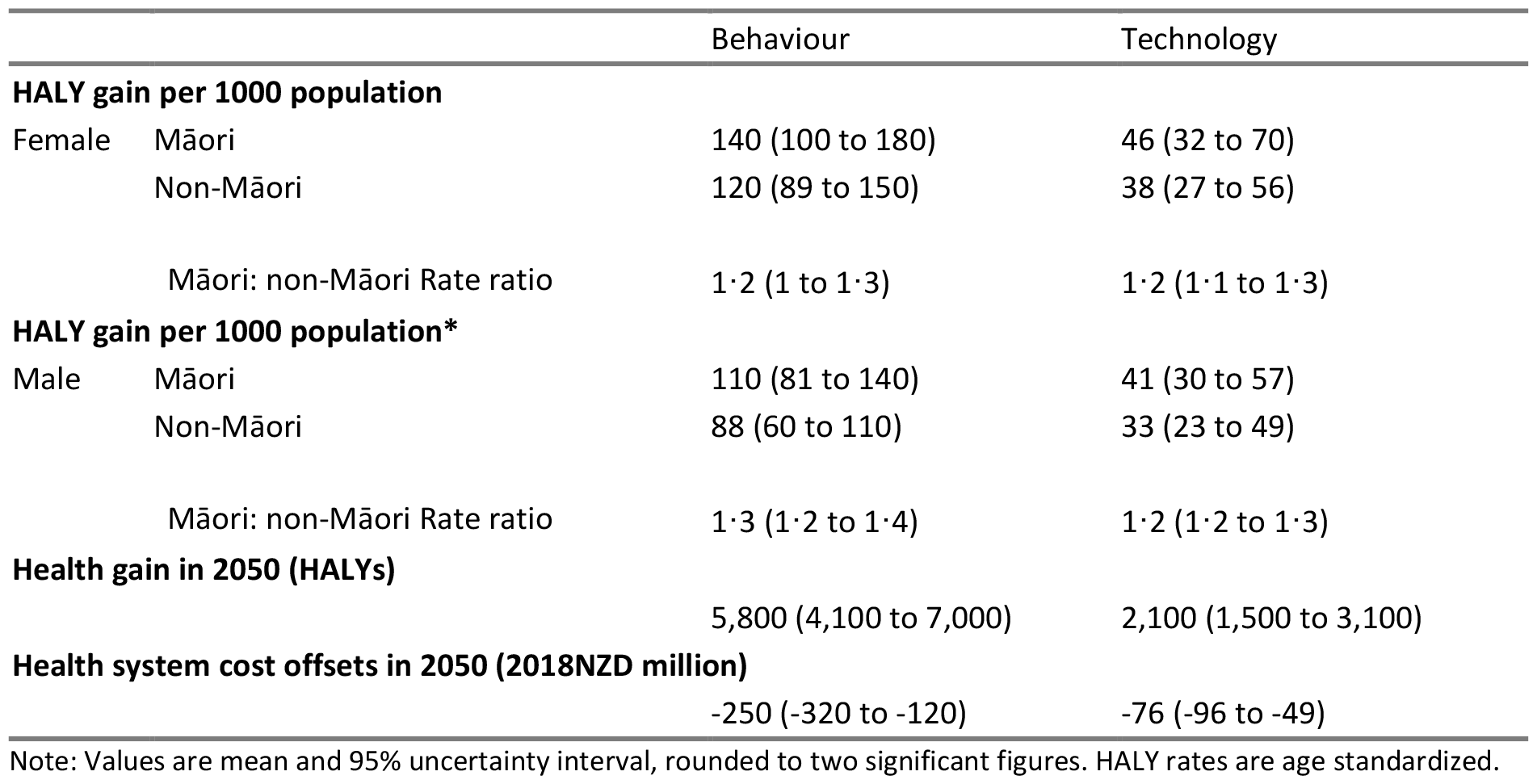
Summary health gains in 2050 for Māori and non-Māori by pathway.

Cumulatively, between 2018 to 2050 the Behaviour pathway would result in 57,000 (95%UI 35,000 to 71,000) HALYs gained and $3200 million savings (95%UI -4,200 to -1,200) in health system costs, while the Technology pathway would result in 22,000 (95%UI 15,000 to 33,000) HALYs gained and $980 million savings (95%UI -1,300 to -530) in health system costs (see SI Figure 5)

Both CCC transport pathways showed changes in healthy life expectancy (see Table 7 in SI). For Māori, gains in added days of healthy life are larger in the younger cohorts (aged 0-4 and 30-34 in 2018) and in the Behaviour pathway. For example, in the 0-4 years cohort the life expectancy gains would reduce the gap between Māori and non-Māori healthy life expectancies by about 0·5% in the behaviour pathway and 0·2% in the technology pathway. However, in the 60-64 year old cohort healthy life expectancy gains, while reduced for everyone, are slightly larger for non-Māori, meaning life expectancy inequities increase slightly.

### Equity scenarios

Figure 4 tests assumptions related to how equitably the pathways are implemented (proportional effect, equitable effect and cycling equity) and the impact of the current inequitable distribution of morbidity and mortality for Māori in Aotearoa on the modelled impacts of the pathways (equal mortality and/or morbidity). Almost all these equity scenarios result in greater health gains for Māori. For example, the cycling equity analysis shows that in the Behaviour pathway if cycling prevalence becomes more gender and ethnicity equitable as mode share increases, the HALY gain for Māori women doubles compared to base case, increasing the rate ratio to 2·6 (95%UI 2.0-3.1). The equal morbidity and mortality scenarios show the modelled health gains that Māori would achieve if they were not penalised in the model by their higher burden of co-morbidities and lower life expectancy. For example, in the Behaviour pathway the rate ratio of health gains for Māori men compared to non-Māori men increases from 1·2 (95%UI 1·2-1·3) in the base case to 2.0 (95%UI 1·9-2·2) with the same level of background morbidity and mortality as non-Māori men.

**Figure 4.**
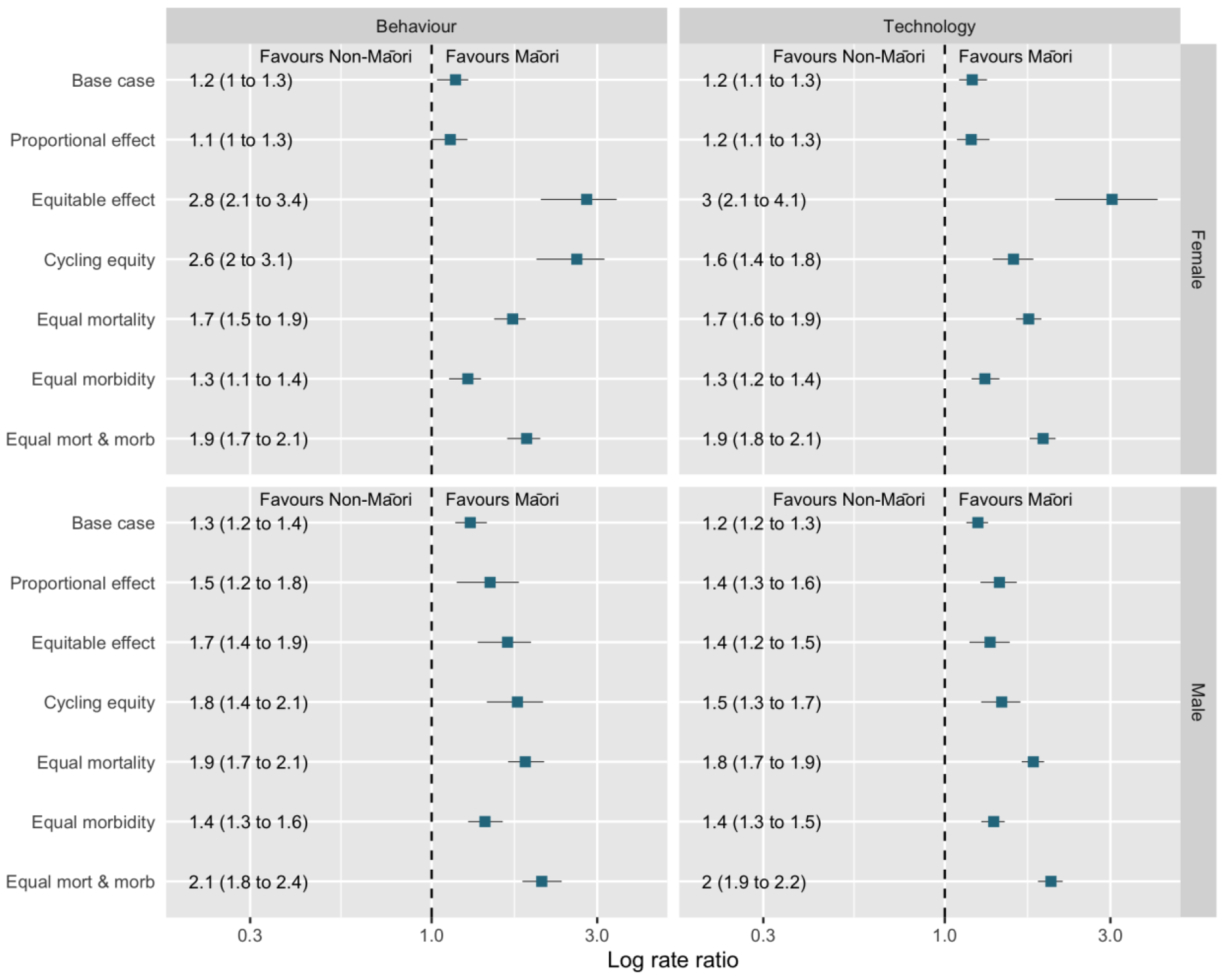
Rate ratios of modelled HALY gains for Māori: non-Māori in 2050 by gender and decarbonisation pathway across selected equity scenario. Base case is the main analysis from Table 4. Explanation of the equity scenarios in Table 1. Full results underpinning this figure in SI Table 8.

### Risk factors and health conditions

Table 5 outlines where modelled health gains are derived from in 2050 by ethnicity and gender. In the Behaviour pathway most of the health gains come through increasing physical activity (74%). However, this varies by ethnicity and gender. For example, 30% of the HALY gains for Māori men come from reductions in transport injury compared to 14% for non-Māori men. In the Technology pathway health benefits are relatively evenly distributed between air pollution and physical activity risk factors, again with comparatively large contribution (30%) coming from injury reductions for Māori men.

**Table 5.**
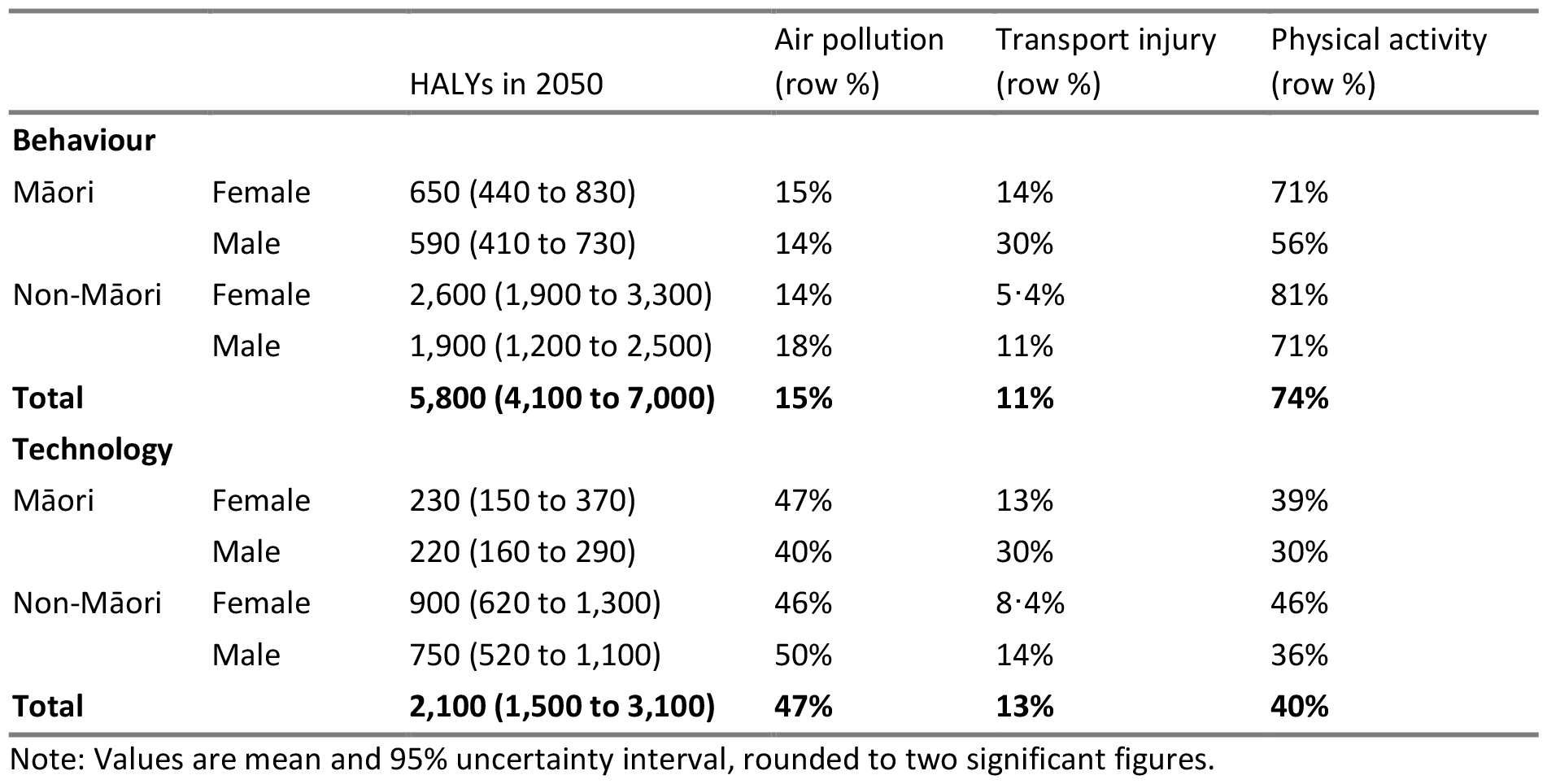
HALY attribution by risk factor in 2050 by gender, ethnicity and decarbonisation pathway.

The SI includes modelled changes in specific health conditions under the pathways. For both pathways most health gains occur through reductions in CVD and diabetes. There was no change in the prevalence of overweight in either pathway but some reductions in the prevalence of obesity in the Behaviour pathway. There were complex patterns for transport injuries; with reductions in vehicle injuries but increases in cycling injuries, all of which varied by gender, ethnicity and time.

Transport injuries increase in the Behaviour pathway, principally due to increases in cycling injury. For Māori there is an increase in transport injury up to the mid-2030s which then decreases, for non-Māori transport injury increases and remains elevated until 2050. These patterns are highly sensitive to our assumptions about how the pathways are implemented. We undertook a sensitivity analysis using a cycling injury rate that from a higher, more equitable cycling environment; this largely eliminated the increase in cycling injuries seen in the base case.

### Greenhouse gas emissions

Greenhouse gas emissions reduce significantly under both pathways compared to BAU (see Table 13 in SI). The total emissions reduction was quite similar in both pathways in 2050, with total emissions reducing from 11000mkg CO2eq to 2,800 (95%UI 2,400 to 3,300) for the Behaviour pathway and 2,600 (95%UI 2,100 to 3,300) in the Technology pathway. Most emission reduction was through tailpipe emissions falling. Sensitivity analyses (full results in SI) of lower ecar uptake show both an erosion of GHGe reductions and a 24% reduction in health gains in the ‘Technology scenario’. Sensitivity analyses around dietary emissions had minimal impacts on GHGe.

## Discussion

### Summary

The results show that decarbonisation of transport may contribute to health equity for Indigenous Peoples in Aotearoa. Both Technology and Behaviour pathways, which represent the CCC’s most contrasting views of transport system changes, could potentially result in greater health gains for Māori compared to non-Māori. The Behaviour pathway has larger modelled health gain, predominately through increased physical activity. Existing inequities in healthy life expectancy between Māori and non-Māori also reduce in both pathways, but more so for the Behaviour pathway. Scenario modelling on how equitably the scenarios are implemented show that impacts on equity are highly dependent on the implementation of the pathways in practice as well as the magnitude of baseline inequities in transport and health.

### Policy and practice implications

This research adds to a substantial body of work confirming potential health benefits of decarbonisation,^4-7^ and the emerging literature examining equity impacts.^12^ The health benefits of the Behaviour pathway are comparable to other accepted public health interventions. For example, the overall health benefits (HALY gains per 1000 population) of the Behaviour pathway sit at a midpoint between tobacco control interventions of 10% tax increases and tobacco free generation (see SI Figure 8).^36^ While specific comparisons for Māori are limited, the scale of healthy life expectancy gains from the Behaviour pathway is around 20-30% of the gains that full tobacco eradication might result in.^37^

The modelling suggests there is potential for decarbonisation to reduce life expectancy inequities between Māori and non-Māori. The equity scenarios we modelled show that the way pathways are implemented has significant impacts on the magnitude of change in life expectancy. For example, moving towards more equitable transport mode distribution by gender and ethnicity in the Behaviour pathway doubles the HALY gain for Māori females compared to the base case and increases the HALY gain for Māori males by up to a quarter. We did not model a scenario whereby Māori have lower uptake of the interventions than non-Māori. However, this is entirely plausible, given the structural inequities that systematically marginalise Māori and limit access to society’s goods and services.^38^ Inequitable implementation of the pathways would reduce health gain for Māori and erode potential contribution of decarbonisation to reductions in life expectancy differences between Māori and non-Māori. Thus, the design and implementation of specific policies within the pathways is fundamental to achieving any reduction in inequities.^39^ This is likely to pose a significant challenge to the transport sector, which has only just started to think about equity in planning and policy.^40^ Moreover in Aotearoa, the transport sector has, until relatively recently, actively pursued policies that discriminate against Māori.^41^

The pathways modelled represent the CCC’s view about potential futures at a certain point in time. In contrast, other Government agencies in Aotearoa have less optimistic views about ecar uptake.^33,42^ In comparison to the UK CCC pathways, the Aotearoa CCC pathways are more optimistic on cycling (cycling VKT is 3 to 13 times higher per week compared with the UK Widespread Engagement and Balanced pathways) and more pessimistic on walking (walking VKT per week is about half as much as the UK Widespread Engagement and Balanced pathways).^4^ The CCC pathways have also not fully explored the potential for mode shift through changes in urban form, public transport and walking for transport.^43^ All the pathways outlined by the CCC represent a conventional view of the transport system which perpetuates the system of car dependence and the inequity that is fundamental to it.^44,45^ In contrast, Indigenous scholars are increasingly suggesting a much more radical approach to planetary health is needed to create systems that enable people to thrive.^46,47^ Finally, what happens in practice may diverge from hypothetical pathways, meaning assessing equity impacts needs to be an ongoing process.

### Strengths and limitations

This is the first model we are aware of that has examined the impact of proposed decarbonisation policy on health equity as well as population health. We examined how the pathways impact on relative and absolute measures of inequity for Indigenous and non-Indigenous people in Aotearoa. Additionally, we undertook a range of analyses that focus on equity within the pathways and make explicit how pre-existing health inequity limits potential health gain for Māori. The model includes 18 different conditions, uses the latest international and local evidence around exposure-response relationships,^20-22^ and includes a wide range of GHGe, including food-related emissions from changes in transport.

This model represents a relatively conservative view of the health gains that could be achieved. Firstly, the model included household travel only, thus excluding the potential gains in air pollution, injury and physical inactivity from decarbonising commercial transport. Secondly, some pathways to health, e.g., noise exposure, were not possible to include in the model. Thirdly, due to data limitations, some health outcomes were either not able to be included at all (e.g. physical activity-dementia^48^ and air pollution-diabetes^49^) or only a very small proportion of the total burden of the condition could be included (e.g. asthma and depression hospitalisations only). Finally, the HALE estimates did not include changes in BMI to avoid double counting of health benefits (many of which are common with physical activity). Other research has shown large changes in HALYs as a result of relatively modest changes in mean BMI.^50^ Finally, the reductions in GHGe would also have health impacts as a result of averted climate change, which we do not account for.^51^

While this model represents a significant advance in examining inequities for Māori compared to non-Māori, we were unable to examine other ethnic groupings (e.g., Pacific peoples) and other important axes of transport-health inequity, such as socioeconomic position and disability, because adequate data were not available.

The model does not account for other policy or technology changes that are occurring in transport. For example, gradual improvements in fleet vehicle efficiency which may reduce vehicle NOx and GHGe and changing weight and size distribution of vehicles which may increase injury and GHGe and make active transport less attractive in urban areas. These will likely all influence decarbonisation pathways as well as population health and health equity and would be fruitful areas for future modelling.

## Conclusion

This work provides evidence that decarbonisation policy has the potential to contribute significantly to health equity between Indigenous and non-Indigenous peoples in Aotearoa. However, the magnitude, and potentially the direction, of impacts on equity depend on the approach to decarbonisation and how equitably the policies to deliver this approach are implemented.

## Data Availability

Data used in this model are not owned by the authors and need to be requested from the relevant institutions that provided them.

## Acknowledgments

- The organisations and individuals who have provided us with data for the model: Ministry of Transport, Ministry of Health, Te Whatu Ora-Health New Zealand, Sport New Zealand, Statistics New Zealand, Waka Kotahi, Emission Impossible and the HAPINZ3 team, Anna Davies, June Atkinson and Giorgi Kvizhinadze.
- Our project advisory group and international advisors
- Cristina Cleghorn for commenting on the draft paper
- Linda Cobiac created the central multistate lifetable model structure in the R programming language while previously employed at Griffith University and we also acknowledge methods assumptions and input data from previous multistate lifetable modelling developed in the BODE^3^ research programme at the University of Otago.

## Author contributions

Caroline Shaw: conceptualization, funding acquisition, methodology, project administration, supervision, writing-original draft. Anja Mizdrak: conceptualization, funding acquisition, software methodology, writing-review and editing. Ryan Gage: formal analysis, methodology, software, writing-review and editing. Melissa McLeod: conceptualization, funding acquisition, methodology, writing-review and editing. Rhys Jones: funding acquisition, writing-review and editing. Alistair Woodward: funding acquisition, writing-review and editing. Linda Cobiac: software, methodology, investigation, visualization, writing-review and editing.

