## Supplementary material for "Policy approaches to decarbonising the transport sector in Aotearoa/NZ: Equity, health and health system impacts"

Contents

Pathways ... 2
Selecting which CCC pathways to model ... 2
Adapting CCC model parameters to our model ... 4
Model parameter inputs ... 8
Methods ... 11
Model input summary ... 12
Sensitivity analyses ... 16
Results ... 18
Equity analyses ... 19
Risk factors ... 22
Diseases and conditions ... 22
Greenhouse gas emissions ... 26
Sensitivity analyses ... 28
Comparisons with other public health interventions ... 30
References ... 31

**Pathways**

The CCC pathway information and input parameters were obtained from publicly available information here.[1] For each pathway the CCC presents information on their assumptions of household annual VKT by mode out to 2050 alongside detailed information on vehicle fleet makeup over the same time frame.

Selecting which CCC pathways to model

The key parameters relevant to our model from the seven pathways developed by the CCC are outlined in Table 1 below. We selected two of the pathways to include in this paper; "Further behaviour change" and "Further technology change". These two were selected as they represented the most diverse perspectives of the Commission about potential pathways to decarbonised household travel. Further behaviour change had the combination of highest levels of VKT reduction, greatest mode shift towards cycling and walking and lowest electrification level of the light fleet. In contrast "Further technology change" had the combination of greatest electrification of the light fleet, lowest mode shift and VKT reduction.

**Table 1 VKT and EV proportion of fleet for CCC pathways in 2050**

| Pathway name | Demonstration path | Headwinds | Further behaviour change* | Further technology change* | Tailwinds | Alt path A | Alt path B |
|---|---|---|---|---|---|---|---|
| **Household person-kilometres travelled by mode (million km) in 2050** | | | | | | | |
| Pedestrian | 1386·0 | 1224·9 | 1398·9 | 1224·9 | 1398·9 | 1398·9 | 1224·9 |
| Cyclist | 3054·3 | 1041·6 | 4581·8 | 1041·6 | 4581·8 | 4581·8 | 1041·6 |
| Micro-mobility | 1·3 | 1·3 | 1·2 | 1·3 | 1·2 | 1·2 | 1·3 |
| Local train | 4747·2 | 3754·6 | 5190·2 | 3754·6 | 5190·2 | 5190·2 | 3754·6 |
| Local bus | 4071·4 | 3242·6 | 4431·0 | 3242·6 | 4431·0 | 4431·0 | 3242·6 |
| Local ferry | 190·5 | 124·1 | 232·4 | 124·1 | 232·4 | 232·4 | 124·1 |
| Motorcycle | 287·6 | 287·6 | 260·1 | 287·6 | 260·1 | 260·1 | 287·6 |
| Taxi / vehicle share | 16215·0 | 16215·0 | 14662·2 | 16215·0 | 14662·2 | 14662·2 | 16215·0 |
| Light vehicle passenger | 12340·7 | 12943·5 | 9910·7 | 12943·5 | 9910·7 | 9910·7 | 12943·5 |
| Light vehicle driver | 22895·5 | 26354·4 | 18278·5 | 26354·4 | 18278·5 | 18278·5 | 26354·4 |
| Total | 65189·5 | 65189·5 | 58946·9 | 65189·5 | 58946·9 | 58946·9 | 65189·5 |
| **EV proportion of vehicle kilometres travelled (%) in 2050** | | | | | | | |
| Light passenger vehicle | 97% | 92% | 89% | 100% | 98% | 92% | 100% |
| Light commercial vehicle | 89% | 81% | 80% | 93% | 91% | 86% | 93% |
| Motorcycle | 71% | 65% | 61% | 74% | 70% | 65% | 74% |
| Medium truck | 88% | 80% | 81% | 89% | 88% | 85% | 89% |
| Heavy truck | 84% | 76% | 72% | 87% | 78% | 82% | 87% |
| Bus | 93% | 87% | 85% | 94% | 94% | 91% | 94% |
| Light | 95% | 89% | 87% | 98% | 96% | 91% | 98% |
| Heavy | 88% | 81% | 81% | 89% | 88% | 86% | 89% |
| All | 95% | 89% | 86% | 98% | 96% | 90% | 98% |

*Modelled in paper

Adapting CCC model parameters to our model

This section outlines how the information from the CCC two pathways was modified to create an input file for our model.

Table 2 outlines where the specific input parameter information was obtained from and Tables 3 and 4 outline the adaptations needed to the CCC parameters to create the final input file for each pathway for our model.

**Table 2 Files used to obtain input parameter information**

| File name | Version date | Data sheets/rows used |
|---|---|---|
| ENZ scenarios dataset for 2021 final advice | 22 June 2021 | Sheets: Further behaviour change and further technology change<br>Lines: 365-445 |
| ENZ assumptions and inputs for 2021 final advice | 14 June 2021 | Sheet: Transport<br>Line 258: Table called: BEV share of EV uptake |

.

**Table 3 Overall input parameters CCC modelling and our model**

| | CCC model | Our model | Differences | Adaptations |
|---|---|---|---|---|
| Household VKT | Spreadsheets provide household VKT by mode 2013-2050. Not stated where data sourced from. | Baseline VKT estimates per person by mode by age, sex and Māori/non-Māori were extracted from analysis of 2015-2018 NZ Household travel survey. | • CCC VKT totals overall and for specific modes for the 2015-2018 period (baseline for the modelling) were different to the VKT from the HH travel survey.<br>• CCC VKT estimates were by household not per person.<br>• CCC VKT estimates took population growth into account. | We derived the annual percentage change in per person travel, by mode, compared with baseline (2018) levels, from the reported *Household person-kilometres travelled by mode*, adjusting for the projected changes in New Zealand population [2] and assuming a constant household size. |
| Light fleet composition | Spreadsheets provide EV proportion of vehicle numbers from 1990 -2050 for full range of vehicle types, estimates of road vehicle kilometres-travelled by vehicle type and engine type from 2001-2050 and BEV share of EV uptake (remainder is PHEV) 2018-2050. Not stated where data sourced from. | Waka Kotahi NZ Transport Agency fleet distribution data for the years 2015-18.[3] | Nil | We assumed the hybrid proportion of the electric vehicle market would linearly reduce to zero over 10 years. |

**Table 4 Mode specific adaptations for CCC input parameters**

| CCC model: VKT by mode | CCC model: engine type parameter for this mode | Our model mode/type | Changes made to harmonise these differences and create an input file for our model and rationale for these. |
|---|---|---|---|
| Cyclist | Assumed all bikes and bike ownership models had negligible/no emissions in their evidence review.[4] | We broke cycling VKT into 4 categories:<br>• Regular cyclist (own bike)<br>• Regular cyclist (bike share)<br>• Ebike cyclist (own bike)<br>• Ebike cyclist (bike share) | For the 2015-2018 time period we assumed the following cycling VKT distribution: Regular cyclists (own bike) 95%, regular cyclists (bike share) 0·1%, Ebike cyclist (own bike) 4·8%, Ebike cyclist (bike share) 0·1%. This assumption is based on researcher knowledge that there were few ebikes or bike share schemes in Aotearoa in 2015-18 (there was no survey information available about the VKT distribution or trips of ebike vs regular bike or models of ownership).<br><br>We then assumed that by 2035 the cycling VKT distribution will have stabilised at: Regular cyclists (own bike) 45%, regular cyclists (bike share) 1%, Ebike cyclist (own bike) 50%, Ebike cyclist (bike share) 4%. This distribution remains the same to 2050. These numbers are based on the assumptions that:<br>• Ebikes become dominant in NZ because of the weather, terrain and low density of cities. Observational studies suggest they are 8-50% of bikes used for commuting currently[5], imports are on an upwards trajectory,[6] and about 6% of the population report using an ebike in 2022, up from 3% in 2018 (personal communication Carol Christie, Waka Kotahi)<br>• Shared schemes remain relatively niche due to their lack of reliability and the lack of 'fit' with people's needs for carrying things (e.g. estimates of shared vs private bike use in Brisbane in 2019 showed only 8% of cycles observed were from a share scheme[7])<br>• Shared schemes are only available in larger urban areas. |
| Micro-mobility (defined in CCC report as escooters and eskateboards)<br><br>Note: the HTS baseline data for our model had no micromobility – so we 'reallocated' VKT from cycling to this category. | Assumed negligible or no emissions from this mode, irrespective of whether they are privately owned or shared.[4] | We broke micromobility VKT into 2 categories:<br>• e-scooter (own)<br>• e-scooter (shared) | For the 2015-2018 time period we assumed the following micromobility VKT distribution: Escooter private 20%, Escooter shared 80%. This assumption is based on researcher knowledge that there were shared escooters in NZ in 2018 and very few private ones (there was no objective information was available to inform this assumption).<br><br>We then assumed that by 2025 the micromobility VKT distribution will have stabilised at: e-scooter private 70%, e-scooter shared 30%. This distribution remains the same to 2050. These numbers are based on the assumptions that:<br>• Shared ebike use may be a pathway to private purchase eg An observational study in Brisbane in 2019 noted that shared escooter use declined from 55% of scooter use to 36% in a 6 month period.[7]<br>• People also report being more likely to want a private escooter[8]<br>• Private escooter imports in NZ are increasing.[6] |
| Walking | N/A | Pedestrian walking | No change needed |
| Motorcycle | Assumptions around transition of motorcycles from ICE to electric for each pathway were contained in the CCC spreadsheets. | We broke motorcycle VKT into 2 categories:<br>• Electric motorcycle<br>• ICE motorcycle | No change needed |
| CCC had light vehicle VKT broken into these 3 categories:<br>• Taxi / vehicle share<br>• Light vehicle passenger | In the CCC information:<br>• Conventional hybrids were grouped with ICE vehicles[4]. | We broke light vehicle VKT into the following groups:<br>• Motorvehicle diesel | We:<br>• Grouped up the VKT from the three light vehicle categories in the CCC information. This may result in some underestimates for air pollution and GHGe as we can't account for any 'dead head' trips in the 'share' category. No information was |

| CCC model: VKT by mode | CCC model: engine type parameter for this mode | Our model mode/type | Changes made to harmonise these differences and create an input file for our model and rationale for these. |
|---|---|---|---|
| • Light vehicle driver | • It was not possible to separate out the proportion of ICE light vehicles that were diesel vs petrol powered.<br>• Estimates of the proportion of the light fleet that were electric and plugin hybrid were available for 2018-2050. | • Motorvehicle ev<br>• Motorvehicle hev (conventional hybrid)<br>• Motorvehicle petrol<br>• Motorvehicle phev (plugin hybrid)<br>We did not distinguish between passenger, driver or share trips in our model. | provided in the CCC main report about what this share category consisted of e.g. taxis, ride hails, individuals car pooling so it was hard to know how to treat it.<br>• Used current % of light vehicle fleet that is diesel and assumed these were phased out at the same rate as petrol cars from the pathways.<br>• We assumed the hev and phev would be phased out over 10 years. (The impact on the results of this assumption will be minimal since the proportions of hev and phev are tiny.) |
| Local bus | Assumptions around transition of buses from ICE to electric for each pathway were contained in the CCC spreadsheets. | We broke bus VKT into the following groups:<br>• Public transport ebus<br>• Public transport bus | No change needed |
| Local train | It wasn't entirely clear what the CCC assumed about passenger rail as fuels for rail are combined with fuel for road in the table "Liquid fossil fuel consumption by mode (PJ)" and this includes rail for freight as well as local rail. | Public transport train | We assumed all local passenger rail was electric. This is not entirely accurate currently but will be shortly and the low VKT associated with this mode meant it would not make much difference to the health or emissions components of the model. |

NB: Local ferry VKT was less than <0·4% of total VKT of two pathways modelled and has few health implications so we did not include it

Model parameter inputs

**Figure 1 Change in annual km travelled by mode, sex and ethnicity: Behaviour pathway**

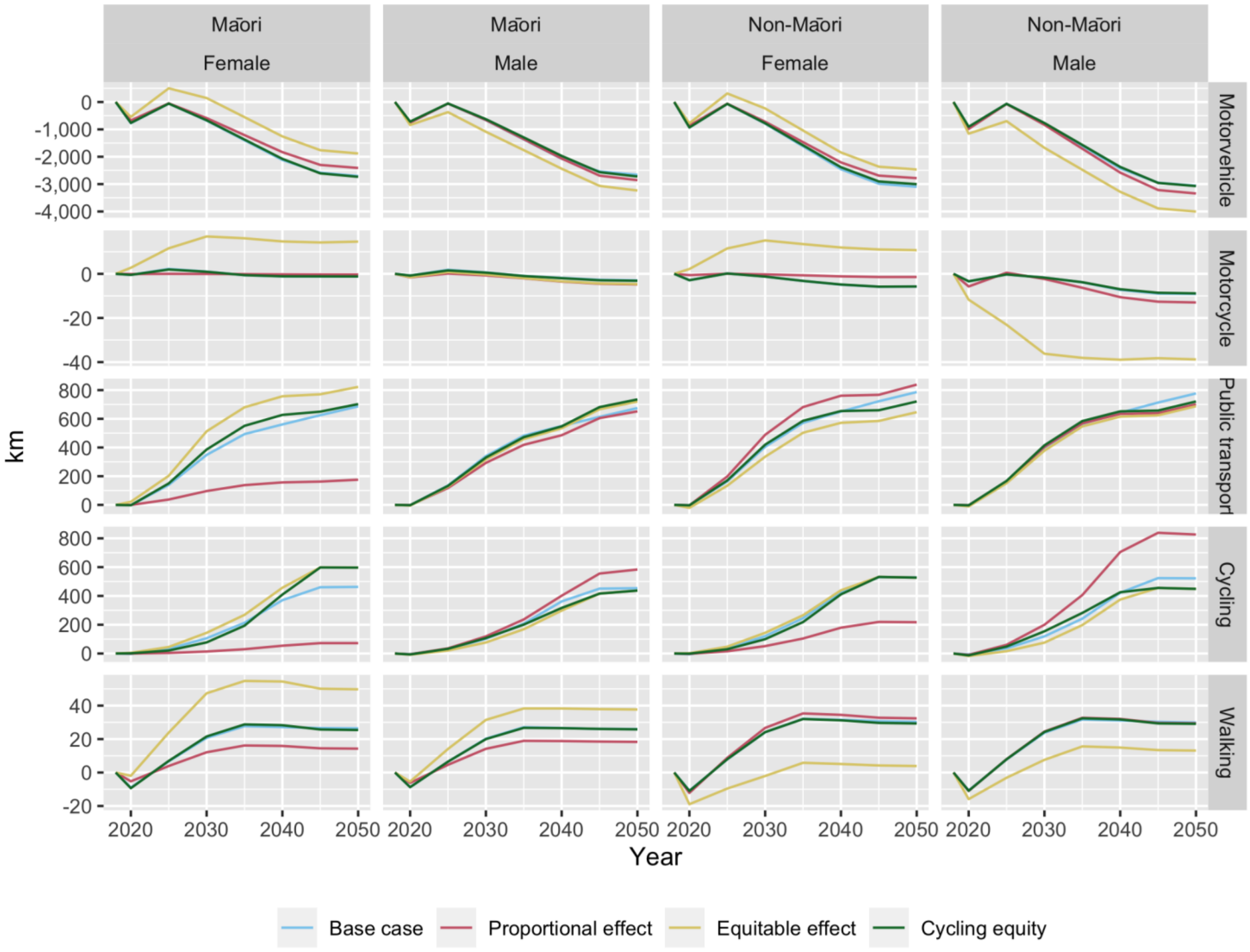

**Figure 2 Change in annual km travelled by mode, sex and ethnicity: Technology pathway**

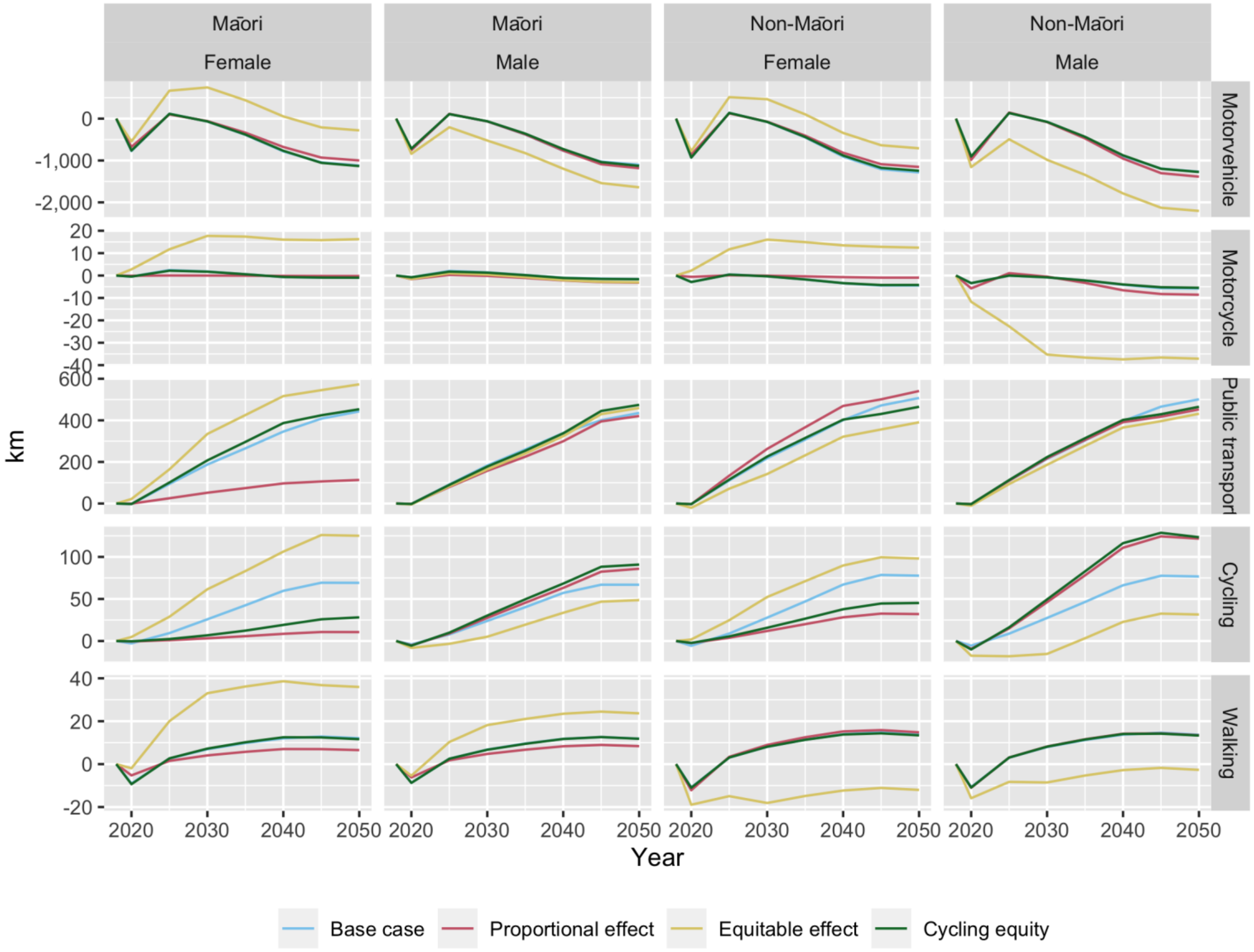

**Methods**

**Figure 3 Model overview**

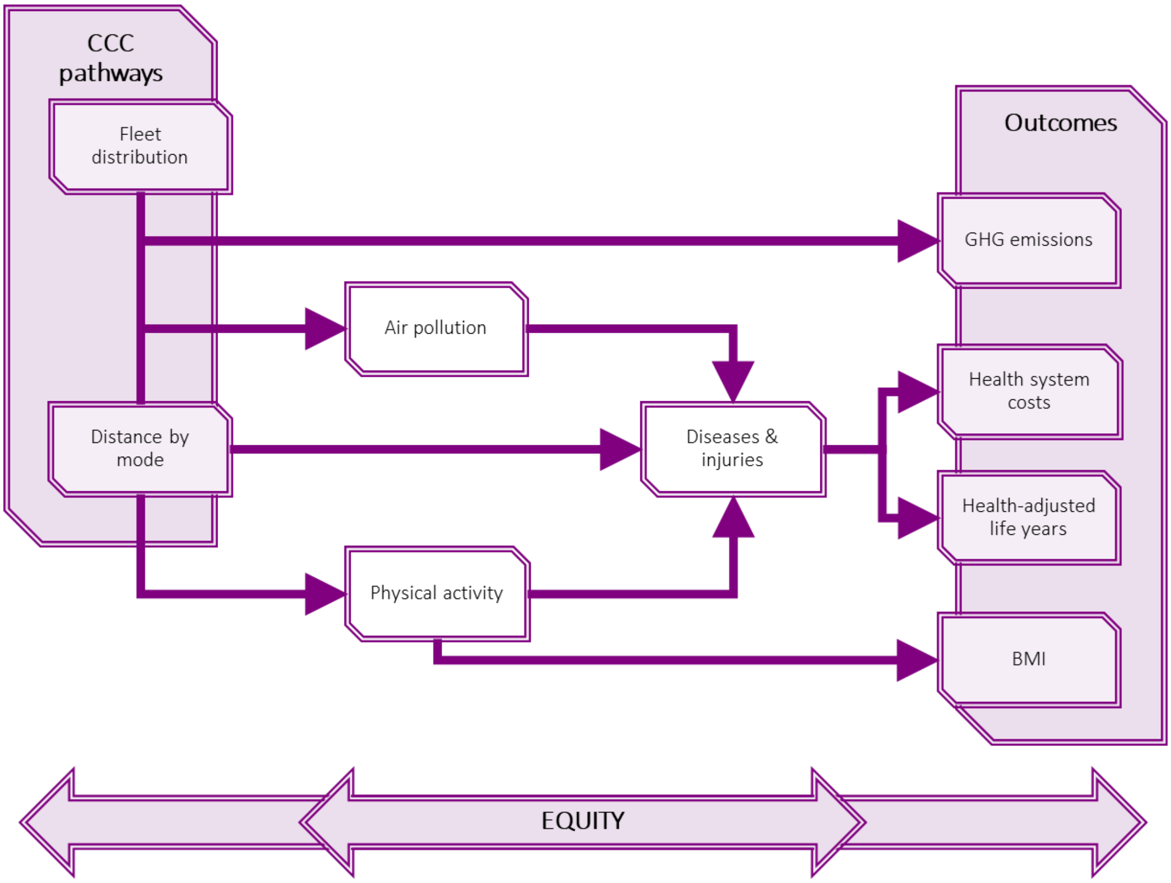


CCC: Climate Change Commission, GHG: Greenhouse gas; BMI: Body Mass Index

Model input summary

**Table 5 Model input summary: data sources, disaggregation and uncertainty distribution**

| Input category | Description of source(s) | Disaggregation | Uncertainty distribution |
|---|---|---|---|
| **Population demographics** | | | |
| Population | Estimated resident population count based on 2018 Census from Statistics NZ. Future cohorts of 0-4 year olds based on 2016 median national population projections.[2] | Age, sex, ethnicity | – |
| All-cause mortality | NZ Period Lifetables 2017-2019 from Statistics NZ[9] | Age, sex, ethnicity | – |
| All-cause/ background morbidity | Based on the NZ Burden of Disease Study[10-12] | Age, sex, ethnicity | – |
| **Transport data** | | | |
| Distance by mode | Total per person weekly kilometres travelled from the NZ Household Travel Survey 2015-2018.[13] Modes included were pedestrian, cyclist, motor vehicle, public transport. | Age, sex, ethnicity | – |
| Fleet distribution | Waka Kotahi NZ Transport Agency fleet distribution data for the years 2015-18.[3] | Not available | – |
| **Diseases and injuries** | | | |
| Chronic* conditions | Incidence, prevalence, and mortality rates were extracted from Ministry of Health National Collections. Disease rates were smoothed and processed using disbayes[14,15] to obtain a coherent set of disease parameters complete with estimates for case fatality. Conditions included: ischaemic heart disease, Stroke, tracheal, bronchus and lung cancer, Head and neck cancer, Liver cancer, uterine cancer, Colon and rectal cancer, Breast cancer, Stomach cancer, Leukaemia, Multiple myeloma, Type 2 diabetes. | Age, sex, ethnicity | – |
| Acute* conditions | Incidence and mortality rates were obtained from National Collections. Conditions included: Pedestrian Road injury, Cyclist road injury, Motorbike road injury, Motor vehicle road injury, Asthma, major depressive disorder (incidence only) | Age, sex, ethnicity | – |
| Cause-specific disability rates | Rates of disability were derived by dividing years lived with disability from the Global Burden of Disease | Sex, ethnicity | – |

| Input category | Description of source(s) | Disaggregation | Uncertainty distribution |
|---|---|---|---|
| | study by the number of prevalent cases (chronic conditions) or incident cases (acute conditions).[16] | | |
| **Physical activity** | | | |
| Exposure | Estimates of moderate/vigorous physical activity weekly METS created using 2017/2018 NZ Health Survey data[3] adjusted to remove occupational PA using Sport NZ Active NZ Survey 2019/20.[17] | Age, sex, ethnicity | – |
| Relative risks | Relative risks from meta-analyses.[18,19] | Sex | Lognormal |
| METS and speeds of active modes | *Walking:* 2·0 MET (based on NZ HTS data on speed of walking in NZ 4·4km hr[20]).<br>*Conventional cycling:* 5·8 MET (based on 2011 compendium for Physical activity - 01011 bicycling, to/from work, self-selected pace.) Speed assumption 16km/hr (lower bound of the 01011 MET value description).[21]<br>*Ebiking:* 4·97 METS (reduced METS of conventional cycling by 0·83, based on meta-analysis[22]). Speed 19km/hr (upper bound of the PA compendium 01011 MET value).[21]<br>*Escootering:* 1·5 MET (based on 2011 compendium for Physical activity - 09701 miscellaneous standing. There is no data available on METS of escootering (as far as we are aware), however some research suggests it is less than walking and/or cycling[23,24] so we used a lower value than walking). Assumed speed of 8·8km/hr (double that of walking).<br>**Note:** all MET values noted here have been adjusted down 1 MET for baseline physical activity. | – | Lognormal |
| Energy compensation assumption (i.e., what proportion of energy expended in additional active travel is then consumed in food - which is relevant to BMI and food-related GHGe calculations) | 57% (UI 19-96%) based on local and international literature.[25,26] | - | |
| **Air pollution** | | | |
| Exposure | $NO_2$ and $PM_{2·5}$ exposure from HAPINZ study[27] Proportion of pollution from modes associated with household travel: 29·4% for $NO_2$ and 5·1% for $PM_{2·5}$.[28] | – | Lognormal |
| Relative risks | Relative risks of disease based on $NO_2$ and $PM_{2·5}$ exposure from NZ specific cohort study. We included the following relationships that were significantly associated with relevant health outcomes:<br>- $NO_2$ exposure and asthma morbidity (all ages) and mortality (ages 30+) | – | Lognormal |

| Input category | Description of source(s) | Disaggregation | Uncertainty distribution |
|---|---|---|---|
| | - $NO_2$ exposure and stroke morbidity (relative risk applied to stroke incidence for ages 30+)<br>- $PM_{2 \cdot 5}$ exposure and ischaemic heart disease and stroke morbidity (relative risk applied to IHD incidence for ages 30+)[29] | | |
| Air pollution emissions (motorised modes) | $NO_2$ and $PM_{2 \cdot 5}$ emissions per kilometre estimated from light vehicle and bus fleet data in the NZ Vehicle Emissions Prediction Model.[30] | – | Lognormal |
| **Injury** | | | |
| Safety-in-numbers | Coefficients for motor vehicles/motorbikes, cyclists and pedestrians from meta-analysis of transport studies[31] | – | Beta |
| Injury proportions (victim/striker) | The breakdown of injury rates based on the mode of travel of the victim and striker (if any) was based on hospitalisation data from the Ministry of Health National Collections. | Sex, ethnicity | |
| **BMI** | | | |
| Exposure | Mean and standard deviation of BMI from the NZ Health Survey 2016/17 - 2018/19.[32] | Age, sex, ethnicity | – |
| Resting metabolic rate | Mean resting metabolic rate from the NZ Health Survey 2017/18.[3] | Age, sex, ethnicity | – |
| Effect of energy expenditure on body weight | Dose response in kJ/day/kg from Hall et al. [33] | – | Normal |
| **Costs** | | | |
| Health-system costs | Disease-specific costs separated into incidence, prevalence (chronic conditions only), last 6 months of life and background costs.[34] Costs for this work were derived from Ministry of Health National Collections data for financial years 2015/16 to 2017/18, Age groupings used varied by disease. | Age, sex | Lognormal |
| **GHGe** | | | |
| Vehicle -tailpipe (includes electricity for ecars) | Equivalent $CO_2$ emissions per kilometre travelled by mode (motor vehicle, motor cycle,bus and rail) and fuel type (petrol, diesel, electric, hybrid and plug-in hybrid),[35] adjusted for vehicle occupancy using the NZ Household Travel Survey, where needed.[13] Equivalent $CO_2$ emissions estimated per kilometre travelled by e-bike and e-scooter use estimated per person per km from published estimates,[36] adjusted for NZ electricity generation mix.[37] | - | Lognomal |

| Input category | Description of source(s) | Disaggregation | Uncertainty distribution |
|---|---|---|---|
| Vehicle- other<br>Includes emissions related to:<br>1) Manufacturing/recycling: includes emissions relating to vehicle manufacturing, maintenance and disposal of vehicles,<br>2) Infrastructure: includes emissions related to construction, maintenance and end-of-life management of infrastructure required for vehicle operations, and<br>3) Operational service emissions: emissions related to the additional commuting required to move the vehicle to the area of demand (relevant to shared e-bikes and e-scooters). | Equivalent $CO_2$ emissions per kilometre travelled by mode (motor vehicle, motor cycle,bus, rail, e-bike and e-scooter) and fuel type (petrol, diesel, electric, hybrid and plug-in hybrid),[36] | - | Lognormal |
| Food- related (active modes) | For each active mode we calculated kcal/km required to ‘fuel’ an individual using data from the NZHS[32] and MET/speed data for each mode described above. We then multiplied this by average gGHGe/100kcal of the NZ diet (310) which was calculated from values in Drew.[38] This gave food-related emissions factors (gCO2e/km) of 133 for walking, 94 for conventional cycling, 83 for ebiking, 30 for escootering. | - | Lognormal |

* Conditions/diseases are classified as chronic or acute based on whether they were modelled as a disease with progression beyond one year or as an acute event/exacerbation of a condition.

Sensitivity analyses

**Table 6 Sensitivity analysis inputs and justification**

| Parameter | Sensitivity analysis | Justification |
|---|---|---|
| Electric car uptake | Lower uptake – with 75% of fleet electric by 2050 | Based on NZ Ministry of Transport estimates, which are more conservative than the CCC.[39] |
| Cycling injury rates | 96% reduction in cycling injury incidence rates and 94% reduction in cycling mortality rates, based on comparison with the Netherlands | Analysis of cycling injury rates[16] and distances travelled by cycle in NZ[13] and European countries, where cycling is much more prevalent suggests that the rate of injuries per km travelled in New Zealand are very high (see below) For example, compared with the Netherlands, rates of non-fatal cycling injuries are 24 times higher and rates of fatal injuries are 15 times higher in New Zealand. Moreover, non-Māori males currently predominate in cycling participation[40] and related injuries (our data) in Aotearoa/New Zealand. As our base case modelling amplifies these current transport patterns, we thought that safety in numbers adjustments alone would be unlikely to fully capture the likely changes in injury risk which would occur with more equitable patterns that occur with higher prevalence of cycling.[41] |
| Dietary GHGe | 30% less $CO_2$ intensive than current NZ diet. | We also estimated impacts with a more climate-friendly diet with 30% less $CO_2$ emissions in sensitivity analyses.[38] |

**Figure 4 Annual cycling injuries and deaths relative to km cycled NZ compared to a range of European countries.**

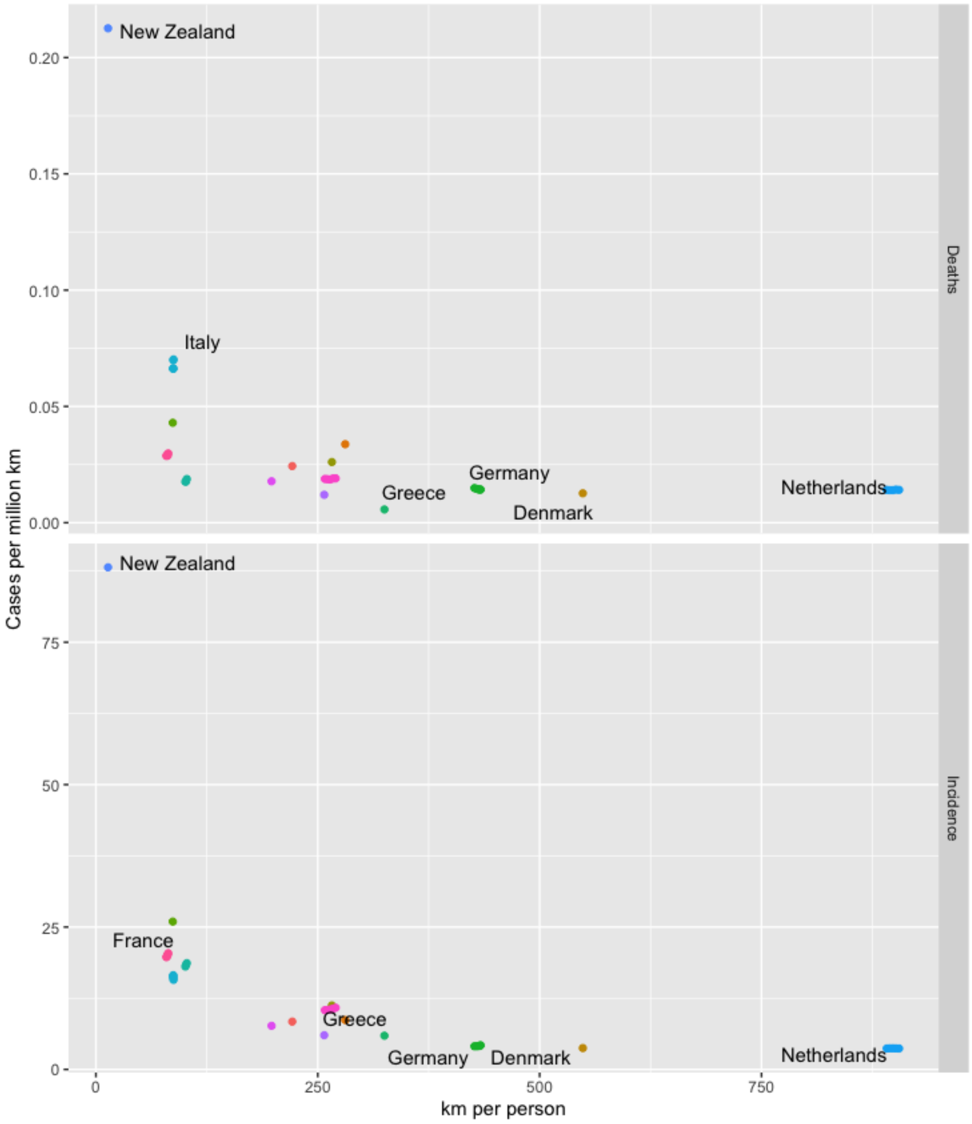


Source: NZ data: data from NZHTS and MOH as part of the model inputs, European data from Castro et al[42] (countries rated as having a 'High' or 'Very high' reliability of data on cycling distance travelled included).

### Results

Overall results

**Figure 5 Population health gain and health system savings by pathway cumulatively to 2050**

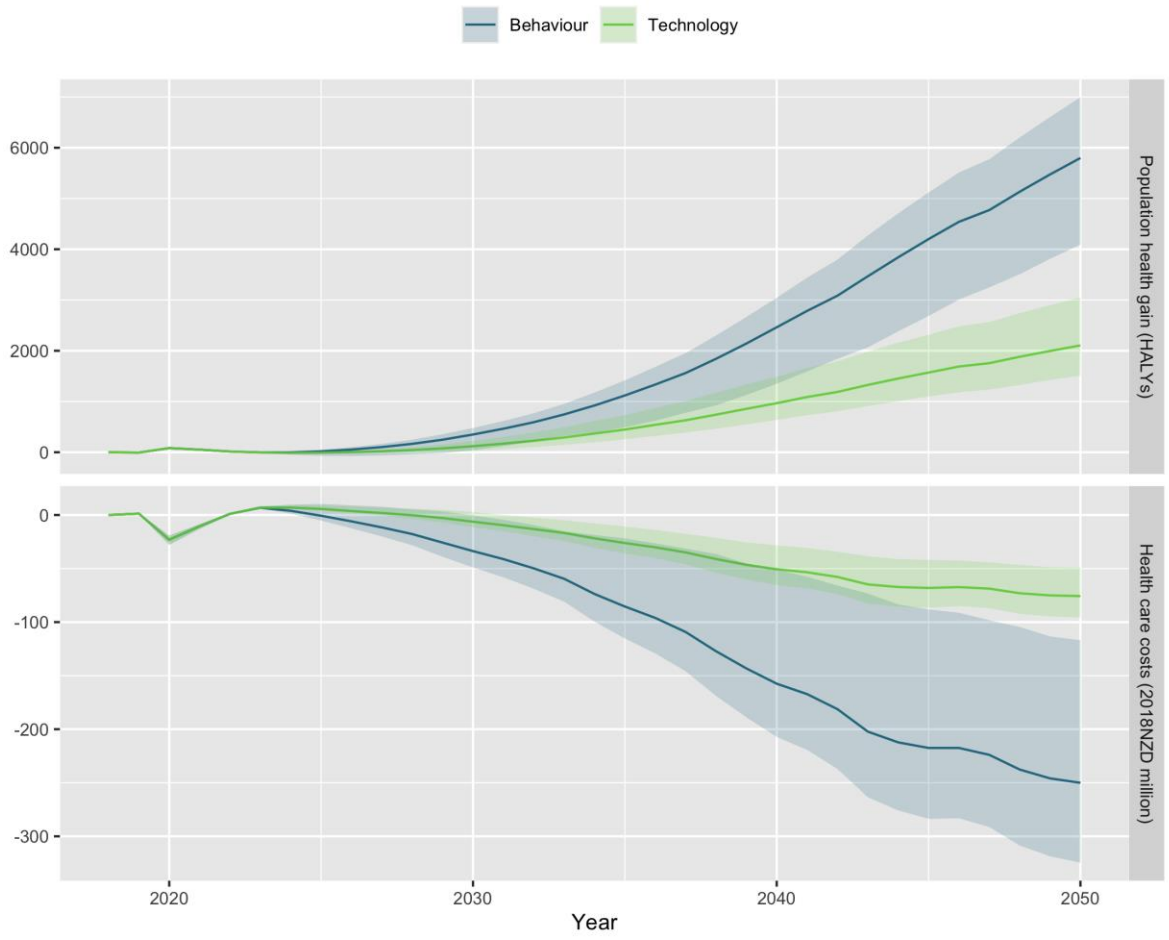


**Table 7 Healthy life expectancy gains by age cohort in 2018, gender and pathway for Māori and non-Māori**

| | Māori | Non-Māori | Difference |
|---|---|---|---|
| **Female: 0-4 years** | | | |
| Baseline | 63 years | 71 years | 8·3 years |
| Behaviour | +69 (51 to 87) days | +54 (40 to 66) days | -0·51% |
| Technology | +23 (16 to 33) days | +17 (12 to 24) days | -0·2% |
| **Female: 30-34 years** | | | |
| Baseline | 36 years | 43 years | 7·3 years |
| Behaviour | +48 (33 to 63) days | +43 (31 to 54) days | -0·2% |
| Technology | +16 (10 to 24) days | +14 (9·7 to 20) days | -0·068% |
| **Female: 60-64 years** | | | |
| Baseline | 14 years | 18 years | 4·8 years |
| Behaviour | +9·6 (6·4 to 13) days | +16 (11 to 20) days | 0·34% |

| | | | |
|---|---|---|---|
| Technology | +3·5 (2·2 to 5·6) days | +5·9 (3·9 to 9·4) days | 0·14% |
| **Male: 0-4 years** | | | |
| Baseline | 61 years | 70 years | 9 years |
| Behaviour | +58 (43 to 72) days | +42 (29 to 53) days | -0·49% |
| Technology | +21 (16 to 28) days | +16 (11 to 22) days | -0·18% |
| **Male: 30-34 years** | | | |
| Baseline | 34 years | 42 years | 7·8 years |
| Behaviour | +37 (26 to 49) days | +30 (20 to 39) days | -0·26% |
| Technology | +14 (9·5 to 19) days | +12 (8 to 18) days | -0·069% |
| **Male: 60-64 years** | | | |
| Baseline | 12 years | 17 years | 4·7 years |
| Behaviour | +5·4 (3·4 to 7·3) days | +7·1 (4·7 to 9·4) days | 0·099% |
| Technology | +2·1 (1·4 to 3·2) days | +3·4 (2·2 to 5·5) days | 0·075% |

Note: Values are mean and 95% uncertainty interval, rounded to two significant figures.

Equity analyses

**Table 8 HALY gain total, and by ethnicity and gender and healthcare cost offsets for equity analyses by pathway**

| | HALY gain per 1000 population | | | | | | Total population health gain in 2050 (HALYs) | Health care cost offsets in 2050 (2018NZD million) |
|---|---|---|---|---|---|---|---|---|
| | Female | | | Male | | | | |
| | Māori | Non-Māori | Rate ratio | Māori | Non-Māori | Rate ratio | | |
| **Behaviour** | | | | | | | | |
| Base case | 140 (100 to 180) | 120 (89 to 150) | 1·2 (1 to 1·3) | 110 (81 to 140) | 88 (60 to 110) | 1·3 (1·2 to 1·4) | 5,800 (4,100 to 7,000) | -250 (-320 to -120) |
| Proportional effect | 73 (53 to 96) | 64 (48 to 82) | 1·1 (1 to 1·3) | 150 (100 to 200) | 100 (69 to 130) | 1·5 (1·2 to 1·8) | 4,700 (3,200 to 5,900) | -230 (-310 to -100) |
| Equitable effect | 280 (170 to 370) | 99 (71 to 120) | 2·8 (2·1 to 3·4) | 140 (97 to 180) | 85 (62 to 100) | 1·7 (1·4 to 1·9) | 5,800 (4,100 to 7,000) | -260 (-330 to -130) |
| Cycling equity | 290 (190 to 380) | 110 (80 to 140) | 2·6 (2 to 3·1) | 130 (90 to 170) | 76 (52 to 95) | 1·8 (1·4 to 2·1) | 5,800 (4,100 to 7,000) | -260 (-340 to -130) |
| Equal mortality | 210 (150 to 260) | 120 (89 to 150) | 1·7 (1·5 to 1·9) | 160 (120 to 210) | 88 (60 to 110) | 1·9 (1·7 to 2·1) | 6,100 (4,300 to 7,400) | -260 (-330 to -120) |
| Equal morbidity | 150 (110 to 200) | 120 (89 to 150) | 1·3 (1·1 to 1·4) | 120 (89 to 160) | 88 (60 to 110) | 1·4 (1·3 to 1·6) | 5,900 (4,200 to 7,100) | -250 (-320 to -120) |
| Equal mort & morb | 230 (160 to 290) | 120 (89 to 150) | 1·9 (1·7 to 2·1) | 180 (130 to 230) | 88 (60 to 110) | 2·1 (1·8 to 2·4) | 6,200 (4,400 to 7,500) | -260 (-330 to -120) |
| Lower injury rate | 140 (110 to 180) | 120 (96 to 150) | 1·2 (1 to 1·3) | 120 (88 to 140) | 92 (71 to 110) | 1·3 (1·2 to 1·4) | 6,100 (5,000 to 7,200) | -270 (-340 to -220) |
| Lower food CO2 | 140 (100 to 180) | 120 (89 to 150) | 1·2 (1 to 1·3) | 110 (81 to 140) | 88 (60 to 110) | 1·3 (1·2 to 1·4) | 5,800 (4,100 to 7,000) | -250 (-320 to -120) |
| MoT EV uptake | 140 (96 to 170) | 120 (86 to 140) | 1·2 (1 to 1·3) | 110 (77 to 140) | 84 (57 to 110) | 1·3 (1·2 to 1·5) | 5,600 (3,900 to 6,700) | -250 (-320 to -110) |
| **Technology** | | | | | | | | |
| Base case | 46 (32 to 70) | 38 (27 to 56) | 1·2 (1·1 to 1·3) | 41 (30 to 57) | 33 (23 to 49) | 1·2 (1·2 to 1·3) | 2,100 (1,500 to 3,100) | -76 (-96 to -49) |
| Proportional effect | 34 (22 to 57) | 28 (19 to 45) | 1·2 (1·1 to 1·3) | 53 (40 to 70) | 37 (26 to 53) | 1·4 (1·3 to 1·6) | 2,000 (1,400 to 2,900) | -74 (-96 to -48) |
| Equitable effect | 86 (53 to 120) | 29 (18 to 46) | 3 (2·1 to 4·1) | 51 (39 to 67) | 38 (29 to 53) | 1·4 (1·2 to 1·5) | 2,200 (1,600 to 3,100) | -81 (-100 to -56) |
| Cycling equity | 50 (34 to 74) | 32 (22 to 49) | 1·6 (1·4 to 1·8) | 52 (38 to 69) | 36 (25 to 52) | 1·5 (1·3 to 1·7) | 2,000 (1,400 to 3,000) | -78 (-100 to -50) |
| Equal mortality | 67 (46 to 100) | 38 (27 to 56) | 1·7 (1·6 to 1·9) | 60 (43 to 83) | 33 (23 to 49) | 1·8 (1·7 to 1·9) | 2,200 (1,600 to 3,200) | -78 (-98 to -50) |
| Equal morbidity | 50 (34 to 76) | 38 (27 to 56) | 1·3 (1·2 to 1·4) | 46 (33 to 63) | 33 (23 to 49) | 1·4 (1·3 to 1·5) | 2,100 (1,500 to 3,100) | -76 (-96 to -49) |

| | | | | | | | | |
|---|---|---|---|---|---|---|---|---|
| Equal mort & morb | 73 (50 to 110) | 38 (27 to 56) | 1·9 (1·8 to 2·1) | 67 (48 to 93) | 33 (23 to 49) | 2 (1·9 to 2·2) | 2,300 (1,600 to 3,300) | -78 (-98 to -50) |
| Lower injury rate | 47 (33 to 71) | 39 (28 to 57) | 1·2 (1·1 to 1·3) | 42 (31 to 58) | 34 (25 to 49) | 1·2 (1·1 to 1·3) | 2,200 (1,600 to 3,100) | -82 (-98 to -68) |
| Lower food CO2 | 46 (32 to 70) | 38 (27 to 56) | 1·2 (1·1 to 1·3) | 41 (30 to 57) | 33 (23 to 49) | 1·2 (1·2 to 1·3) | 2,100 (1,500 to 3,100) | -76 (-96 to -49) |
| MoT EV uptake | 37 (26 to 51) | 32 (23 to 43) | 1·2 (1·1 to 1·3) | 33 (25 to 44) | 27 (19 to 37) | 1·2 (1·2 to 1·3) | 1,600 (1,200 to 2,100) | -71 (-89 to -45) |

Note: Values are mean and 95% uncertainty interval, rounded to two significant figures. HALY rates are age-standardised.

Risk factors

**Table 9 Impact on air pollution ($\mu g/m^3$) in 2050 by gender, ethnicity and pathway**

| | $NO_2$ | $PM_{2\cdot5}$ |
|---|---|---|
| **Baseline values in 2050** | | |
| BAU | 6·8 | 8·4 |
| Behaviour | 4·8 (1·4 to 12) | 8·1 (4·8 to 13) |
| Technology | 4·7 (1·4 to 11) | 8·1 (4·8 to 13) |
| **Difference in 2050** | | |
| Behaviour | -2 (-4·9 to -0·6) | -0·31 (-0·51 to -0·17) |
| Technology | -2·2 (-5·3 to -0·64) | -0·28 (-0·48 to -0·14) |

Note: Values are mean and 95% uncertainty interval, rounded to two significant figures.

**Table 10 Impact on prevalence of physical inactivity (<450 METmins/wk) in 2050 by gender, ethnicity and pathway**

| | **Māori** | | **Non-Māori** | |
|---|---|---|---|---|
| | **Female** | **Male** | **Female** | **Male** |
| **Baseline values in 2050** | | | | |
| BAU | 38 | 23 | 36 | 27 |
| Behaviour | 29 (29 to 29) | 17 (17 to 17) | 26 (26 to 26) | 20 (20 to 20) |
| Technology | 37 (37 to 37) | 22 (22 to 22) | 34 (34 to 34) | 26 (26 to 26) |
| **Difference in 2050** | | | | |
| Behaviour | -9·2 (-9·2 to -9·2) | -6·2 (-6·2 to -6·2) | -10 (-10 to -10) | -7·7 (-7·7 to -7·7) |
| Technology | -1·7 (-1·7 to -1·7) | -1·1 (-1·1 to -1·1) | -1·8 (-1·8 to -1·8) | -1·4 (-1·4 to -1·4) |

Note: Values are mean and 95% uncertainty interval, rounded to two significant figures.

Diseases and conditions

*Diseases*

**Figure 6 Incident cases of disease and injury averted over time under different pathways by gender and ethnicity**

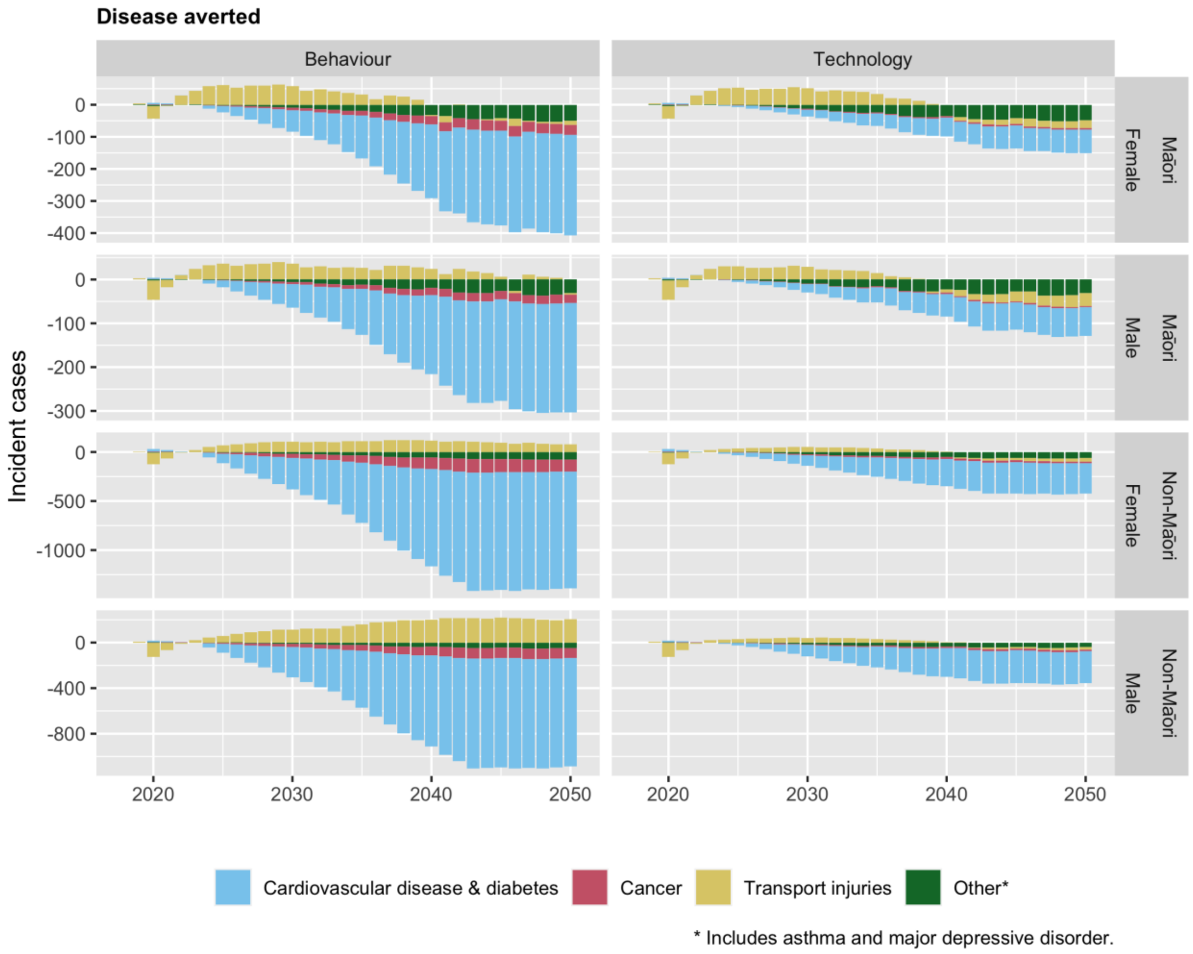
Disease averted
Behaviour
Technology
Incident cases
Māori
Female
Male
Non-Māori
2020
2030
2040
2050
Cardiovascular disease & diabetes
Cancer
Transport injuries
Other*
* Includes asthma and major depressive disorder.

**Table 11 Impact on new cases of disease in 2050 by pathway, gender and ethnicity**

| | Māori | | Non-Māori | |
|---|---|---|---|---|
| | **Female** | **Male** | **Female** | **Male** |
| **Behaviour** | | | | |
| Cardiovascular disease & diabetes: | | | | |
| Diabetes mellitus type 2 | -200 (-280 to -120) | -180 (-260 to -110) | -580 (-800 to -350) | -620 (-870 to -380) |
| Ischemic heart disease | -65 (-100 to -32) | -44 (-69 to -21) | -330 (-510 to -150) | -210 (-320 to -100) |
| Stroke | -45 (-67 to -27) | -21 (-39 to -6·5) | -290 (-420 to -170) | -130 (-230 to -44) |
| Cancer: | | | | |
| Breast cancer | -14 (-21 to -7·5) | | -63 (-91 to -35) | |
| Colon and rectum cancer | -0·87 (-3·7 to 1·8) | -5 (-9·1 to -1·1) | -9·4 (-36 to 16) | -39 (-69 to -12) |
| Head and neck cancer | -2·3 (-5·3 to 1·1) | -4·5 (-12 to 3) | -14 (-32 to 5·5) | -26 (-69 to 19) |
| Liver cancer | -0·45 (-0·96 to 0·074) | -2·5 (-5·1 to 0·15) | -0·83 (-1·7 to -0·01) | -5·3 (-10 to 0·26) |
| Multiple myeloma | -0·63 (-1·9 to 0·65) | -0·5 (-2 to 0·85) | -3·3 (-9·6 to 2·9) | -1·8 (-11 to 6) |
| Stomach cancer | -0·15 (-0·59 to 0·33) | -0·25 (-1·1 to 0·53) | -0·66 (-2·1 to 0·83) | -0·92 (-3·1 to 1·2) |
| Tracheal, bronchus, and lung cancer | -8·4 (-22 to 8) | -6·3 (-18 to 7·7) | -17 (-44 to 14) | -15 (-46 to 20) |
| Uterine cancer | -3·8 (-8·3 to 0·76) | | -14 (-30 to 1·6) | |
| Transport injuries: | | | | |
| Cyclist road injuries | 140 (-0·63 to 1,000) | 160 (-2·9 to 860) | 390 (-3·5 to 2,400) | 500 (-12 to 2,600) |
| Motor vehicle road injuries | -87 (-98 to -76) | -120 (-130 to -100) | -200 (-220 to -170) | -190 (-210 to -160) |
| Motorcyclist road injuries | -67 (-67 to -67) | -47 (-52 to -42) | -110 (-110 to -100) | -100 (-110 to -93) |
| Pedestrian road injuries | 1·6 (-1·6 to 7) | -0·058 (-5 to 8·9) | -5 (-11 to 6·6) | -5·8 (-14 to 8·9) |
| Other: | | | | |
| Asthma | -43 (-120 to -8·8) | -27 (-77 to -6) | -52 (-150 to -10) | -34 (-95 to -7·2) |
| Major depressive disorder | -7·6 (-17 to 0·83) | -3·1 (-8·8 to 2·2) | -23 (-51 to 4·4) | -12 (-31 to 5·5) |
| **Technology** | | | | |
| Cardiovascular disease & diabetes: | | | | |
| Diabetes mellitus type 2 | -36 (-49 to -21) | -32 (-46 to -18) | -100 (-140 to -62) | -110 (-150 to -65) |
| Ischemic heart disease | -21 (-32 to -12) | -21 (-35 to -11) | -100 (-160 to -56) | -99 (-160 to -53) |
| Stroke | -18 (-34 to -8·4) | -12 (-28 to -4·3) | -110 (-210 to -52) | -75 (-170 to -26) |
| Cancer: | | | | |
| Breast cancer | -1·9 (-3·2 to -0·53) | | -8·5 (-14 to -3·1) | |
| Colon and rectum cancer | -0·0038 (-0·55 to 0·53) | -0·67 (-1·5 to 0·078) | -0·26 (-5·6 to 4·8) | -5·7 (-11 to -0·45) |
| Head and neck cancer | -0·4 (-0·96 to 0·19) | -0·8 (-2·1 to 0·53) | -2·4 (-5·8 to 0·96) | -4·8 (-13 to 3·3) |
| Liver cancer | -0·057 (-0·15 to 0·037) | -0·39 (-0·85 to 0·094) | -0·12 (-0·27 to 0·025) | -0·85 (-1·8 to 0·16) |
| Multiple myeloma | -0·1 (-0·33 to 0·13) | -0·074 (-0·35 to 0·17) | -0·58 (-1·7 to 0·54) | -0·31 (-1·9 to 1·1) |
| Stomach cancer | 0·012 (-0·071 to 0·1) | 0·026 (-0·13 to 0·17) | -0·0009 (-0·27 to 0·28) | -0·029 (-0·43 to 0·37) |

| | | | | |
|---|---|---|---|---|
| Tracheal, bronchus, and lung cancer | -1·1 (-3·6 to 1·7) | -0·77 (-2·9 to 1·6) | -2·5 (-7·3 to 2·8) | -2·1 (-7·4 to 4) |
| Uterine cancer | -0·58 (-1·4 to 0·21) | | -2·2 (-5·2 to 0·56) | |
| Transport injuries: | | | | |
| Cyclist road injuries | 44 (-0·2 to 230) | 39 (-1·1 to 160) | 110 (-1·3 to 510) | 110 (-4·8 to 460) |
| Motor vehicle road injuries | -35 (-40 to -30) | -47 (-54 to -41) | -79 (-90 to -69) | -74 (-85 to -65) |
| Motorcyclist road injuries | -35 (-38 to -32) | -23 (-26 to -20) | -66 (-72 to -59) | -60 (-66 to -53) |
| Pedestrian road injuries | 1·7 (0·5 to 3) | 1·4 (-0·38 to 3·5) | -0·66 (-2·9 to 1·9) | -0·52 (-3·3 to 2·7) |
| Other: | | | | |
| Asthma | -47 (-130 to -11) | -30 (-83 to -7) | -57 (-160 to -13) | -37 (-100 to -8·4) |
| Major depressive disorder | -1·5 (-3·1 to 0·073) | -0·55 (-1·6 to 0·38) | -4·2 (-9·7 to 1·2) | -2·2 (-5·7 to 1) |

Note: Values are mean and 95% uncertainty interval, rounded to two significant figures

*Overweight and obesity*

**Table 12 Prevalence of overweight and obesity by pathway, age, gender and ethnicity in 2050**

| | Age group | Overweight | | | | Obese | | | |
|---|---|---|---|---|---|---|---|---|---|
| | | Māori | | Non-Māori | | Māori | | Non-Māori | |
| | | Female | Male | Female | Male | Female | Male | Female | Male |
| **BAU** | 18-34 | 25 | 35 | 28 | 35 | 52 | 41 | 29 | 27 |
| | 35-64 | 26 | 29 | 30 | 38 | 56 | 57 | 39 | 37 |
| | 65+ | 30 | 30 | 35 | 42 | 44 | 57 | 33 | 33 |
| | Total | 26 | 31 | 31 | 38 | 53 | 50 | 35 | 33 |
| **Behaviour** | 18-34 | 25 (25 to 26) | 35 (35 to 36) | 28 (28 to 28) | 34 (34 to 35) | 50 (49 to 52)* | 38 (35 to 40)* | 28 (27 to 29) | 24 (21 to 26)* |
| | 35-64 | 26 (26 to 27) | 30 (29 to 31) | 30 (30 to 30) | 39 (38 to 39) | 55 (54 to 56) | 55 (52 to 57)* | 38 (36 to 39) | 34 (31 to 36)* |
| | 65+ | 30 (30 to 30) | 31 (30 to 32) | 35 (35 to 35) | 42 (42 to 42) | 43 (41 to 44) | 54 (51 to 56)* | 32 (30 to 33) | 30 (27 to 32)* |
| | Total | 26 (26 to 27) | 32 (32 to 33) | 31 (31 to 31) | 38 (38 to 38) | 51 (50 to 53)* | 47 (45 to 50)* | 33 (32 to 35)* | 30 (27 to 33)* |
| **Technology** | 18-34 | 25 (25 to 25) | 35 (35 to 35) | 28 (28 to 28) | 35 (35 to 35) | 52 (51 to 52) | 40 (40 to 41) | 29 (29 to 29) | 26 (26 to 27) |
| | 35-64 | 26 (26 to 26) | 29 (29 to 29) | 30 (30 to 30) | 38 (38 to 38) | 56 (56 to 56) | 57 (56 to 57) | 39 (39 to 39) | 36 (36 to 37) |
| | 65+ | 30 (30 to 30) | 30 (30 to 30) | 35 (35 to 35) | 42 (42 to 42) | 44 (44 to 44) | 56 (56 to 56) | 33 (33 to 33) | 32 (32 to 33) |
| | Total | 26 (26 to 26) | 32 (32 to 32) | 31 (31 to 31) | 38 (38 to 38) | 53 (52 to 53) | 50 (49 to 50) | 35 (34 to 35) | 33 (32 to 33) |

Note: Values are mean and 95% uncertainty interval, rounded to two significant figures. Greater than 1 unit difference between pathway and BAU is starred.

Greenhouse gas emissions

**Table 13 Mean GHGe by source and 95% uncertainty interval by pathway in 2050 (million kg CO2 eq)**

| | Vehicle tailpipe | Vehicle other | Food-related | Total |
|---|---|---|---|---|
| **Baseline values in 2050** | | | | |
| BAU | 9,300 | 1,300 | 38 | **11,000** |
| Behaviour | 1,300 (1,100 to 1,600) | 1,400 (1,100 to 1,800) | 150 (31 to 290) | **2,800 (2,400 to 3,300)** |
| Technology | 850 (600 to 1,200) | 1,700 (1,200 to 2,300) | 55 (12 to 110) | **2,600 (2,100 to 3,300)** |
| **Difference from baseline in 2050** | | | | |
| Behaviour | -7,900 (-11,000 to -5,600) | 91 (-340 to 550) | 110 (22 to 210) | **-7,700 (-11,000 to -5,300)** |
| Technology | -8,400 (-12,000 to -5,900) | 430 (-130 to 1,000) | 17 (3·5 to 34) | **-8,000 (-11,000 to -5,300)** |

Note: Values are mean and 95% uncertainty interval, rounded to two significant figures. Vehicle other includes 1) Manufacturing/recycling includes emissions relating to vehicle manufacturing, maintenance and disposal of vehicles 2) Infrastructure is emissions related to construction, maintenance and end-of-life management of infrastructure required for vehicle operations and 3) Operational service emissions relate to the additional commuting required to move the vehicle to the area of demand (relevant to shared e-bikes and e-scooters). Full results for 'vehicle other' in supplementary material.

**Table 14 Impact on GHGe in 2050 including subcategories of Vehicle-other**

| | Tailpipe | Vehicle -other | Food-related |
|---|---|---|---|

| | | Vehicle | Infrastructure | Operation | |
|---|---|---|---|---|---|
| **Baseline values in 2050** | | | | | |
| BAU | 9,300 | 840 | 430 | 6·7 | 38 |
| Behaviour | 1,300 (1,100 to 1,600) | 1,000 (720 to 1,400) | 340 (260 to 450) | 22 (16 to 28) | 150 (31 to 290) |
| Technology | 850 (600 to 1,200) | 1,300 (880 to 1,900) | 390 (270 to 550) | 14 (9·7 to 19) | 55 (12 to 110) |
| **Difference in 2050** | | | | | |
| Behaviour | -7,900 (-11,000 to -5,600) | 160 (-250 to 590) | -87 (-260 to 62) | 15 (9·2 to 21) | 110 (22 to 210) |
| Technology | -8,400 (-12,000 to -5,900) | 450 (-73 to 1,000) | -36 (-240 to 160) | 7·2 (2·4 to 13) | 17 (3·5 to 34) |

Note: Values are mean and 95% uncertainty interval, rounded to two significant figures.

Sensitivity analyses

*Summary of key GHGe and HALY changes for main sensitivity analyses*

**Table 15 Health gain, healthcare savings and GHGe in 2050 for sensitivity analyses compared to base case**

| | **Total population health gain (HALYs)** | **Health care cost offsets (2018NZD million)** | **Tailpipe GHGe (million kg CO2 eq)** | **Food GHGe (million kg CO2 eq)** |
|---|---|---|---|---|
| **Behaviour** | | | | |
| Base case | 5,800 (4,100 to 7,000) | -250 (-320 to -120) | 1,300 (1,100 to 1,600) | 150 (31 to 290) |
| Lower injury rate | 6,100 (5,000 to 7,200) | -270 (-340 to -220) | 1,300 (1,100 to 1,600) | 150 (31 to 290) |
| Lower ecar uptake | 5,600 (3,900 to 6,700) | -250 (-320 to -110) | 2,000 (1,700 to 2,400) | 150 (31 to 290) |
| Dietary GHGe | 5,800 (4,100 to 7,000) | -250 (-320 to -120) | 1,300 (1,100 to 1,700) | 100 (22 to 200) |
| **Technology** | | | | |
| Base case | 2,100 (1,500 to 3,100) | -76 (-96 to -49) | 850 (600 to 1,200) | 55 (12 to 110) |
| Lower injury rate | 2,200 (1,600 to 3,100) | -82 (-98 to -68) | 850 (600 to 1,200) | 55 (12 to 110) |
| Lower ecar uptake | 1,600 (1,200 to 2,100) | -71 (-89 to -45) | 2,500 (2,100 to 3,000) | 55 (12 to 110) |
| Dietary GHGe | 2,100 (1,500 to 3,100) | -76 (-96 to -49) | 850 (600 to 1,200) | 39 (8·3 to 77) |

Base case is the main analysis. Note: Values are mean and 95% uncertainty interval, rounded to two significant figures. Lower injury rate= cycling injury rates of Netherlands (96% reduction incidence and 93% reduction in mortality), Lower ecar uptake= uptake of 74% by 2050, Dietary GHGe reduction of 30%.

*Injury sensitivity analyses*

**Figure 7 Transport injuries for Behaviour pathway: base case and lower cycling injury rate**

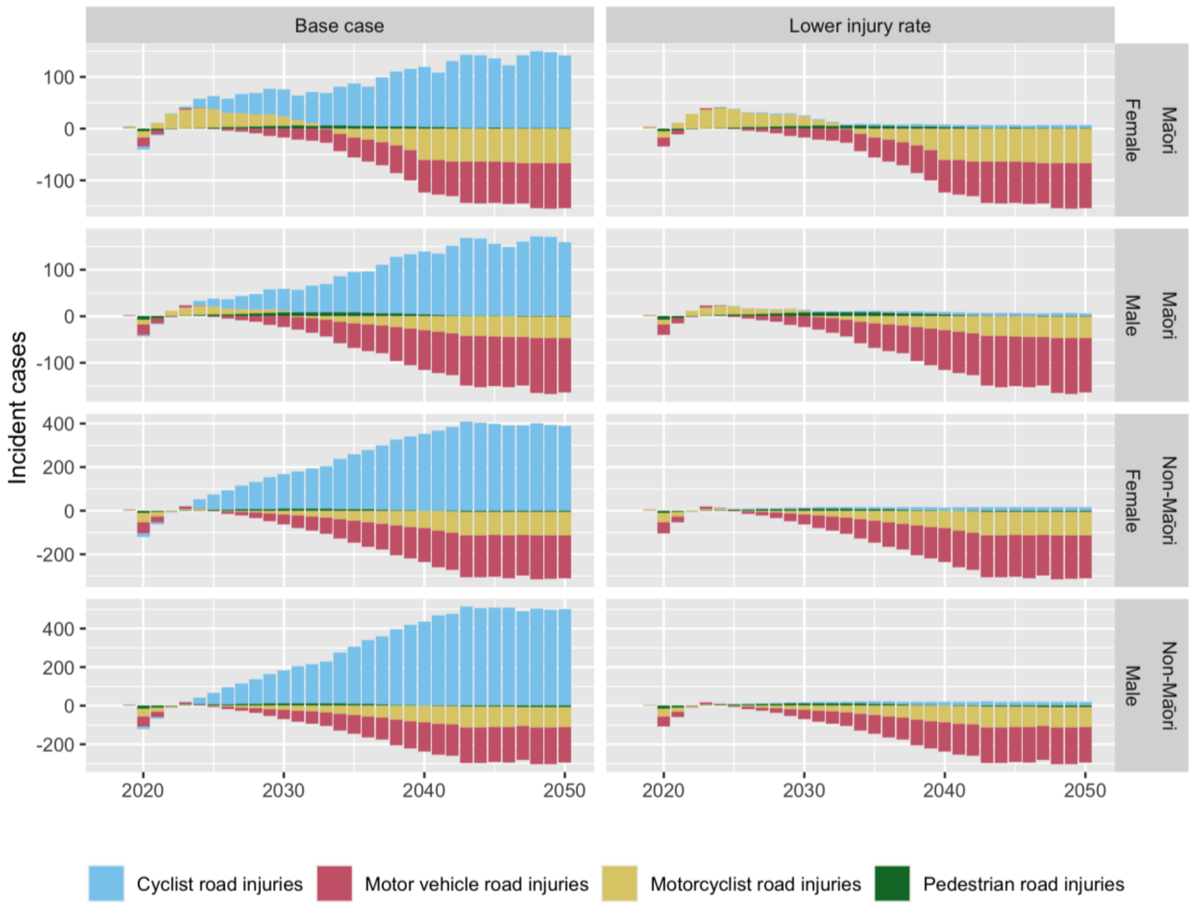

Comparisons with other public health interventions

**Figure 8 Total HALY gain per 1000 population comparison with multistate lifetable literature**

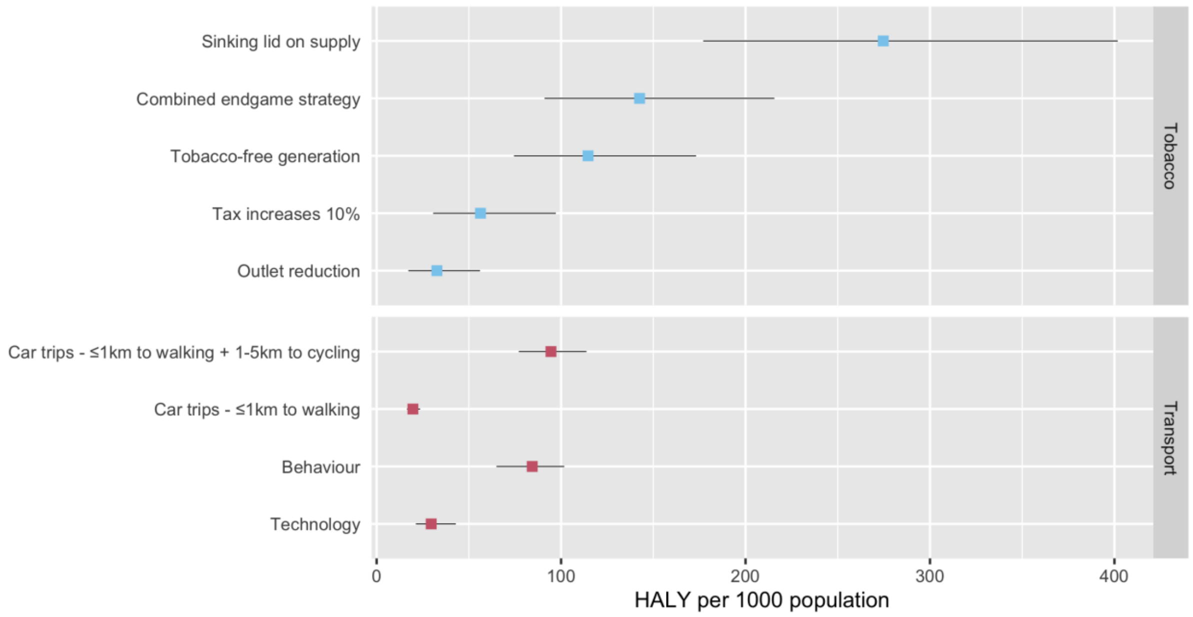


Source: Tobacco analyses: van der Deen et al 2018[43] Car trip switching analysis: Mizdrak et al 2019[44] NB: some differences in methodology so not 100% directly comparable.

**Table 16 Days of HALE gained Behaviour pathway compared to tobacco eradication**

| | **Days of HALE gained in cohort aged 2 in 2011 for tobacco eradication (days of HALE gained)*** | **Cohort aged 0-4 in 2018 days of HALE gained in Behaviour pathway - base case** |
|---|---|---|
| Non-Māori male | 88 (0·24 HALE) | 46 |
| Non-Māori female | 77 (0·21 HALE) | 59 |
| Māori male | 215 (0·59 HALE) | 62 |
| Māori female | 310 (0·85 HALE) | 75 |

*Source: Blakely et al 2020 supplementary table 3[45]

2. Statistics New Zealand. Median projection: National Population Projections: 2016 (base) –2068. Statistics New Zealand 2016.

3. Waka Kotahi NZ Transport Agency. New Zealand vehicle fleet open data sets. 2022. https://nzta.govt.nz/resources/new-zealand-motor-vehicle-register-statistics/new-zealand-vehicle-fleet-open-data-sets/ (accessed 22/11 2022).

4. He Pou a Rangi/Climate Change Commission. Chapter 6: Reducing emissions from transport, buildings and urban form. 2021 Supporting Evidence. Wellington Climate Change Commission; 2021.

5. Lieswyn J, van Mierlo M, Gregory M. Speed surveys of powered transport devices. Report version 10. . ViaStrada Ltd, 2022.

11. Burden of Disease Epidemiology E, and Cost-Effectiveness Programme;. Disease Inputs used for Multi-State Life Table Modelling. In: Burden of Disease Epidemiology E, and Cost-Effectiveness Programme;, editor. Version 1·0 ed; 2019.

12. Blakely T, Cobiac LJ, Cleghorn CL, et al. Health, Health Inequality, and Cost Impacts of Annual Increases in Tobacco Tax: Multistate Life Table Modeling in New Zealand. *PLOS Medicine* 2015; **12**(7): e1001856.

13. Ministry of Transport. New Zealand Household Travel Survey 2015-2017 Methodology Report. Wellington: Ministry of Transport, 2018.

14. Jackson C, Zapata-Diomedi B, Woodcock J. Bayesian multistate modelling of incomplete chronic disease burden data. *Journal of the Royal Statistical Society Series A: Statistics in Society* 2023; **186**(1): 1-19.

15. Jackson C. disbayes: Bayesian Multi-State Modelling of Chronic Disease Burden Data. 1·0·0 ed; 2023. p. R Package.

16. Global Burden of Disease Collaborative Network. Global Burden of Disease Study 2019 (GBD 2019) Results. Seattle, United States: Institute for Health Metrics and Evaluation (IHME); 2020.

17. Sport New Zealand. Active NZ 2018 Participation Report. Wellington: Sport New Zealand, 2019.

27. HAPINZ 3·0. Health Effects Model. 2022.

28. Metcalfe J, Kuschel G. Public Health Risks associated with Transport Emissions in NZ: Part 2 Road Transport Emission Trends. Wellington ESR, 2023.

29. Hales S, Atkinson J, Metcalfe J, Kuschel G, Woodward A. Long term exposure to air pollution, mortality and morbidity in New Zealand: Cohort study. *Science of The Total Environment* 2021; **801**: 149660.

30. Metcalfe J, Peeters S. Vehicle Emissions Prediction Model: VEPM 6·3 technical update report.: Report prepared for Waka Kotahi NZ Transport Agency by Emission Impossible Ltd, 2022.

31. Elvik R, Goel R. Safety-in-numbers: An updated meta-analysis of estimates. *Accident Analysis & Prevention* 2019; **129**: 136-47.

32. Ministry of Health. Methodology Report 2018/19: New Zealand Heath Survey. Wellington: Ministry of Health 2019.

33. Hall KD, Sacks G, Chandramohan D, et al. Quantification of the effect of energy imbalance on bodyweight. *Lancet* 2011; **378**(9793): 826-37.

34. Blakely T, Kvizhinadze G, Atkinson J, Dieleman J, Clarke P. Health system costs for individual and comorbid noncommunicable diseases: An analysis of publicly funded health events from New Zealand. *PLoS Med* 2019; **16**(1): e1002716.

35. Ministry for the Environment. Measuring emissions: A guide for organisations: 2022 detailed guide. Wellington Ministry for the Environment, 2022.

36. International Transport Forum. Good to Go? Assessing the Environmental Performance of New Mobility. Paris: International Transport Forum, 2020.

37. Ministry of Business IE. Energy in New Zealand 2022. Wellington Ministry of Business, Innovation and Employment, 2022.

38. Drew J, Cleghorn C, Macmillan A, Mizdrak A. Healthy and Climate-Friendly Eating Patterns in the New Zealand Context. *Environ Health Perspect* 2020; **128**(1): 17007.

39. Ministry of Transport. Hīkina te Kohupara – Kia mauri ora ai te iwi. Transport Emissions: Pathways to Net Zero by 2050. Wellington Ministry of Transport 2021.

40. Shaw C, Russell M, Keall M, et al. Beyond the bicycle: Seeing the context of the gender gap in cycling. *J Transp Health* 2020; **18**: 100871.

41. Goel R, Goodman A, Aldred R, et al. Cycling behaviour in 17 countries across 6 continents: levels of cycling, who cycles, for what purpose, and how far? *Transport Reviews* 2021; **42**(1): 58-81.

42. Castro A, Kahlmeier S, Gotschi T. Exposure-Adjusted Road Fatality Rates for Cycling and Walking in European Countries. Paris: International Transport Forum, 2018.

43. van der Deen FS, Wilson N, Cleghorn CL, et al. Impact of five tobacco endgame strategies on future smoking prevalence, population health and health system costs: two modelling studies to inform the tobacco endgame. *Tob Control* 2018; **27**(3): 278-86.

44. Mizdrak A, Blakely T, Cleghorn CL, Cobiac LJ. Potential of active transport to improve health, reduce healthcare costs, and reduce greenhouse gas emissions: A modelling study. *PLoS ONE* 2019; **14**(7): e0219316.

45. Blakely T, Cleghorn C, Petrović-van der Deen F, et al. Prospective impact of tobacco eradication and overweight and obesity eradication on future morbidity and health-adjusted life expectancy: simulation study. *J Epidemiol Community Health* 2020; **74**(4): 354-61.
